# How brain networks tic: predicting tic severity through rs-fMRI dynamics in Tourette syndrome

**DOI:** 10.1101/2022.05.24.22275371

**Authors:** Shukti Ramkiran, Tanja Veselinović, Jürgen Dammers, Ravichandran Rajkumar, N. Jon Shah, Irene Neuner

## Abstract

Tourette syndrome (TS) is a neuropsychiatric disorder characterized by motor and phonic tics, which several different theories, such as basal ganglia-thalamo-cortical loop dysfunction and amygdala hypersensitivity, have sought to explain. Previous research has shown dynamic changes in the brain prior to tic onset leading to tics, and this study aims to investigate the contribution of network dynamics to them. For this, we have employed three methods of functional connectivity to resting-state fMRI data - namely the static, the sliding window dynamic and the ICA based estimated dynamic; followed by an examination of the static and dynamic network topological properties. A leave-one-out (LOO-) validated regression model with LASSO regularization was used to identify the key predictors. Most relevant predictors were related to regions belonging to the amygdala-mediated visual social processing network. This is in line with a recently proposed social decision-making dysfunction hypothesis, opening new horizons in understanding tic pathophysiology.

## Introduction

Tourette syndrome (TS) is a complex and chronic neuropsychiatric disorder that presents itself with other comorbidities, such as attention deficit disorder (ADHD) and obsessive-compulsive disorder (OCD), in up to 90% of cases *(1, 2)*. This neurodevelopmental condition is characterized by involuntary movements and vocalizations, known as tics, often present along with emotional and social disturbances *(2)*. There is a growing tendency to understand complex neuropsychiatric disorders such as TS as a network problem rather than as isolated disturbances in specific brain regions *(3)*. For this reason, the interplay within and between large-scale brain networks through the means of brain connectivity analyses is gaining particular interest. Abnormal patterns of static functional connectivity in different brain circuits have been shown in TS *(4–6)*, and several different theories, such as failure of network maturation *(7)*, lack of inhibition *(8)*, basal ganglia circuits dysfunction *(4)* and amygdala dysfunction *(9)*, have tried to explain the different dimensions of the disorder independently. However, several cardinal features, such as the nature of the tics, lifetime prevalence of the disorder and gender disparity, remain unexplained by these different theories. A more recent hypothesis proposes TS to be a disorder of the social decision-making network (SDM) *(10)*, bringing together all the previous models and unexplained cardinal features under one umbrella. This hypothesis puts forth the idea that TS is a disorder of social communication resulting from developmental abnormalities at several levels of the SDM *(10)*.

The gold standard for assessing the severity of tics in patients with TS and other tic disorders is the Yale global tic severity scale (YGTSS). It evaluates the number, frequency, intensity, complexity, and interference of motor and phonic symptoms *(11)*. It is a semi-structured interview followed by a questionnaire which results in five different ratings: total motor tic score, total verbal tic score, total tic score (motor + verbal), overall impairment rating, which reflects the impact of tics on their daily lives and activities and a global severity score. The global score is determined by adding together the total motor, verbal and impairment scores and ranges from 0-100. In clinical practice, the YGTSS is used to track changes in tic behaviour or to monitor treatment outcomes *(12)*. A recent study has shown YGTSS to be robust against the effects of comorbidities such as OCD and ADHD, making it a good choice to investigate the pathophysiology of tics independently of other comorbidities *(13)*.

Dynamic functional connectivity (DFC) refers to the analysis of functional connectivity changes over time. It is effective in capturing temporal variations in spatial connectivity, enabling the identification of different mental states at rest *(14)*. Several studies have shown this method to be useful in identifying transient states of mindfulness and mind-wandering*(14)*, and aberrant transient states in schizophrenia *(15)* and traumatic brain injury *(16)*.

There are multiple approaches for obtaining the DFC from fMRI data. The most common approach is the use of a sliding-window to capture connectivity in short time periods for the whole duration of the scan. This approach is useful in reliably capturing slow dynamics as the frequency is limited by temporal smoothing, i.e., the length of the sliding window. Typically recommended window lengths range from 20 to 30 times the repetition time of the fMRI sequence *(17)*. Another more recent approach to DFC is an estimated form of DFC as opposed to the direct sliding window approach. This is obtained via a group level ICA decomposition of static connectivity, followed by a generalized psycho-physiological interaction model (gPPI) back-projection *(18)*. Although an estimated form, this method allows noise ICA components to be removed, enabling better signal-to-noise ratio, while also making it possible to investigate instantaneous connectivity and faster dynamics. This is especially relevant for TS as patients can experience tics at any point in time, and this sudden onset of tics may be attributed to sudden network switching; it would not be possible to capture this with the standard static network approach. The sudden onset of tics during the resting state has been monitored using a video camera system *(19)*, and changes in brain activation patterns from 2 seconds before tic onset until the actual tic onset have been shown in previous studies *(20)*.

The assessment of network topological properties of the brain allows us to understand the organization and communication strategies employed by the brain. Previous studies on TS have shown disruptions in the balance between local specialization and global integration mechanisms in whole brain-structural networks *(21)* and defects in network maturation, reflected by losses of hub regions in resting-state cortico-basal ganglia functional networks *(6)*. Classical static functional networks show the overall picture of functional organization (division of roles, designation of subnetworks for specialized information processing etc.) in the brain *(18, 22–28)*. However, they lack information about transients or fluctuations in network organization. To bridge this gap, there is growing interest in the temporal network organization or the dynamic graph theory approach *(29)*. This approach offers temporal equivalents of the static graph metrics, and applying it in conjunction with DFC enables the characterization of functional network dynamics. Being a relatively new technique, this study is the first of its kind to investigate its importance in TS.

With converging evidence showing TS as a network disorder and the need to obtain a complete picture of tic pathophysiology, the investigation of network dynamics seems to be a crucial step. Therefore, in this study, we have examined the ability of direct and indirect dynamic network metrics to predict tic severity.

## Results

### Data scope

Imaging data comprising structural MRI, resting-state fMRI, task fMRI and diffusion MRI were acquired from 36 adult TS patients at a 1.5T Siemens scanner as described in *(20)*. Data quality was inspected based on completeness of demographic information, susceptibility and coverage artefacts and length of the scrubbing vector (i.e. no. of valid scans > 50%), and poor-quality data were excluded from further analyses. Hereafter, resting-state fMRI data from 17 adult TS patients were preprocessed using the standard preprocessing procedures described in the Methods section and corrected for artefacts. The patient demographic information and distribution of the YGTSS can be found in **Table 1**.

**Table 1.** Demographic information.

| Age range | Gender | Medication | YGTSS | Comorbidities |
| --- | --- | --- | --- | --- |
| 41-45 | f | - | 33 |  |
| 36-40 | m | - | 68 |  |
| 21-25 | m | 10 mg ESC | 40 |  |
| 21-25 | f | - | 57 |  |
| 41-45 | m | 200 mg TIA | 59 |  |
| 56-60 | f | 40 mg CIT, 400 mg CBZ | 63 | OCD |
| 46-50 | f | - | 46 |  |
| 46-50 | m | - | 54 |  |
| 26-30 | m | - | 17 |  |
| 36-40 | m | 200 mg AMS, 50 mg TIA | 80 | OCD |
| 21-25 | m | - | 2 |  |
| 26-30 | m | - | 63 | OCD, ADHD |
| 21-25 | m | 20 mg CIT, 200 mg TIA | 66 | OCD |
| 26-30 | m | 50 mg TRIM | 44 |  |
| 21-25 | f | 10 mg ARI, 20 mg FLX | 27 |  |
| 16-20 | m | 80 mg ZPR | 60 |  |
| 21-25 | m | - | 62 |  |

CIT – CITALOPRAM, CBZ – CARBAMAZEPINE, PIM – PIMOZIDE, MPH – METHYLPHENIDATE, TIA – TIAPRIDE, AMS – AMISULPIRIDE, TRIM – TRIMIPRAMINE, FLX – FLUOXETINE, ZPR – ZIPRASIDONE, ARI – ARIPIRAZOLE, ESC - ESCITALOPRAM
OCD – OBSESSIVE-COMPULSIVE DISORDER, ADHD – ATTENTION DEFICIT HYPERACTIVITY DISORDER

### Connectivity estimation

The brain was divided into 105 regions of interest (ROIs) covering the cortical and subcortical areas (based on the Harvard-Oxford cortical and subcortical atlases), and the average time series of each ROI was computed. Three types of functional connectivity were computed from each ROI time series, namely:

- The static functional connectivity (sFC), which was obtained by applying a weighted GLM model (bivariate correlation with hrf weighting) to pairs of ROIs.
- The sliding window dynamic connectivity (dSW), which was obtained by temporal decomposition of the ROI time series into sliding windows of length 100s and overlap 75s followed by the computation of bivariate Pearson’s correlation in each sliding window.
- The ICA based dynamic functional connectivity (dICA), which was obtained by group ICA decomposition of the time series of all the subjects followed by gPPI back projection of the independent components to obtain the ROI BOLD responses. The data were decomposed into 20 ICs, and temporal smoothing was applied for 10s. The connectivity ICs arranged in the order of increasing kurtosis can be found in **Figure 1**. The top 5 ICs in the figure were removed before back-projection and dynamic functional connectivity estimation (ICs with kurtosis < 3 were removed; images with low spatial kurtosis yield noise components by identifying uniformly distributed or global connectivity).

**Figure 1:**
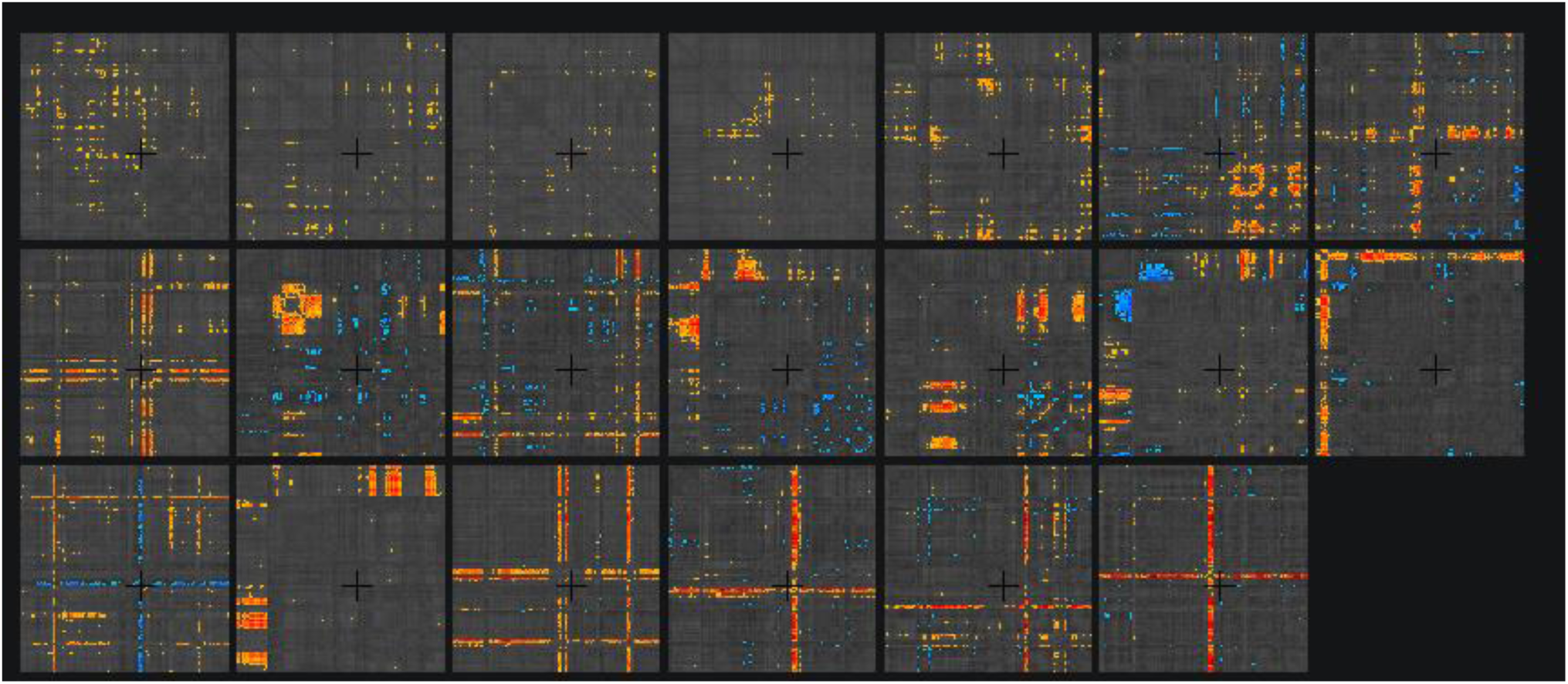
Spatial properties of ICs obtained. The matrices are fully connected, but only connections with z-score > 2 have been displayed. The first five ICs with low spatial kurtosis (<3) were removed before back projection and estimation of the dynamic functional connectivity.

### Calculation of network measures

The connectivity estimates were used to investigate network properties in the brain. Classical static network theory was applied to the static functional connectivity, and a newer dynamic network theory was applied to the dynamic functional connectivity. Both approaches used binarized adjacency matrices, and the binarization threshold was chosen as 0.45 for the correlation coefficient between two ROIs. Thus connections >0.45 or <-0.45 were retained as 1s and others were converted to 0s. This threshold was chosen to retain the maximum number of connections while at the same time obtaining a small world distribution (maximum separation between a random graph and a regular lattice), after inspecting the variable threshold explorer in CONN v20.b. The static network distribution, along with the comparison of regular versus a random graph for different thresholds, can be found in **Figure 2**. To maintain consistency, the same threshold was used for obtaining the dynamic networks. The static and dynamic network measures average path length, clustering coefficient/temporal correlation coefficient, betweenness centrality and small worldness were calculated for each ROI for each subject. The mean and standard deviation of the network measures for each subject can be found in **Figure 3**.

**Figure 2:**
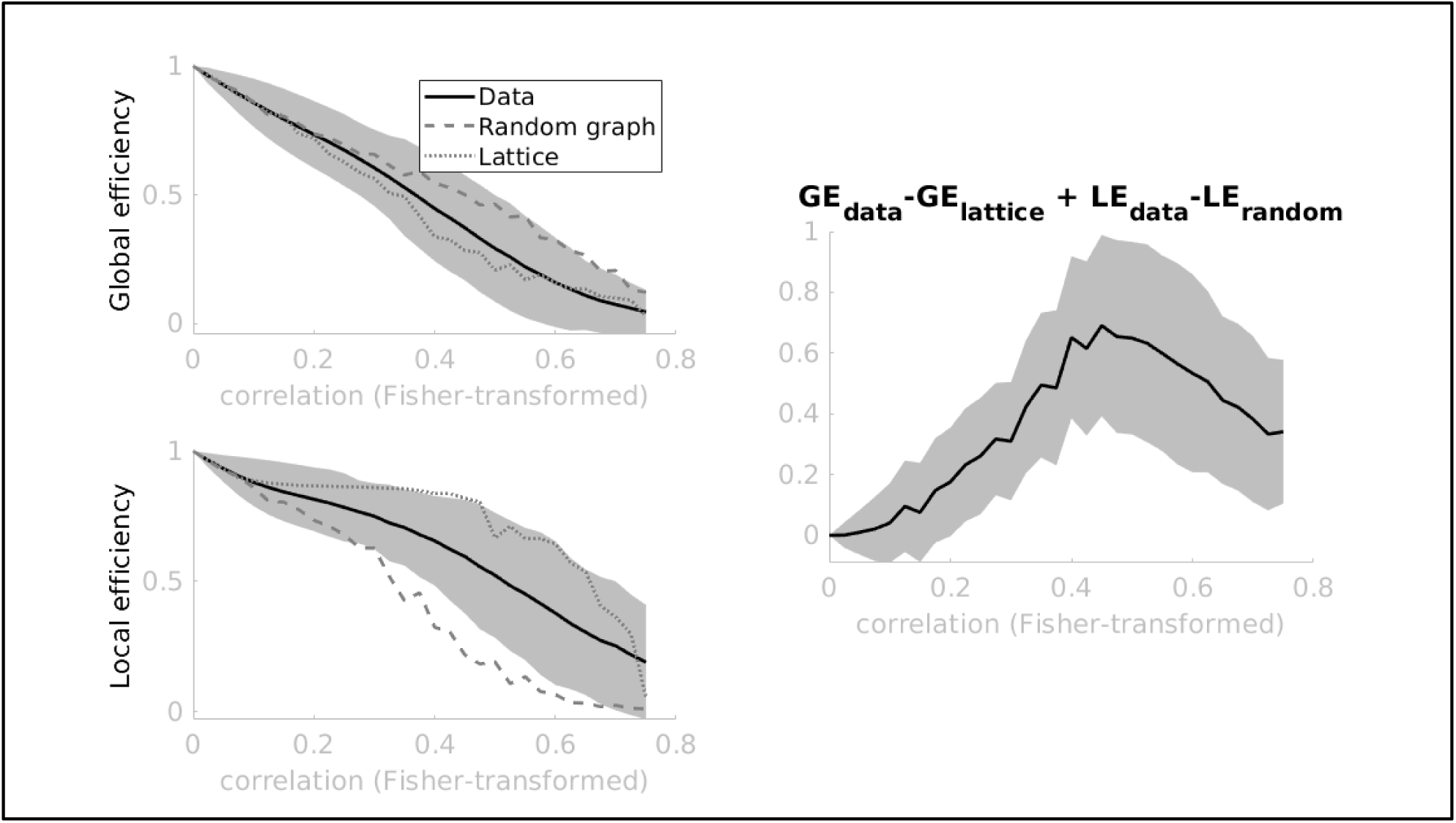
Threshold determination for static network. Variable threshold explorer in CONN v20.b. As we can see, the separation between a random graph and lattice can be best observed between the threshold values 0.4 to 0.6. A value of 0.45 was chosen to retain as many connections as possible while maintaining optimal separation.

**Figure 3:**
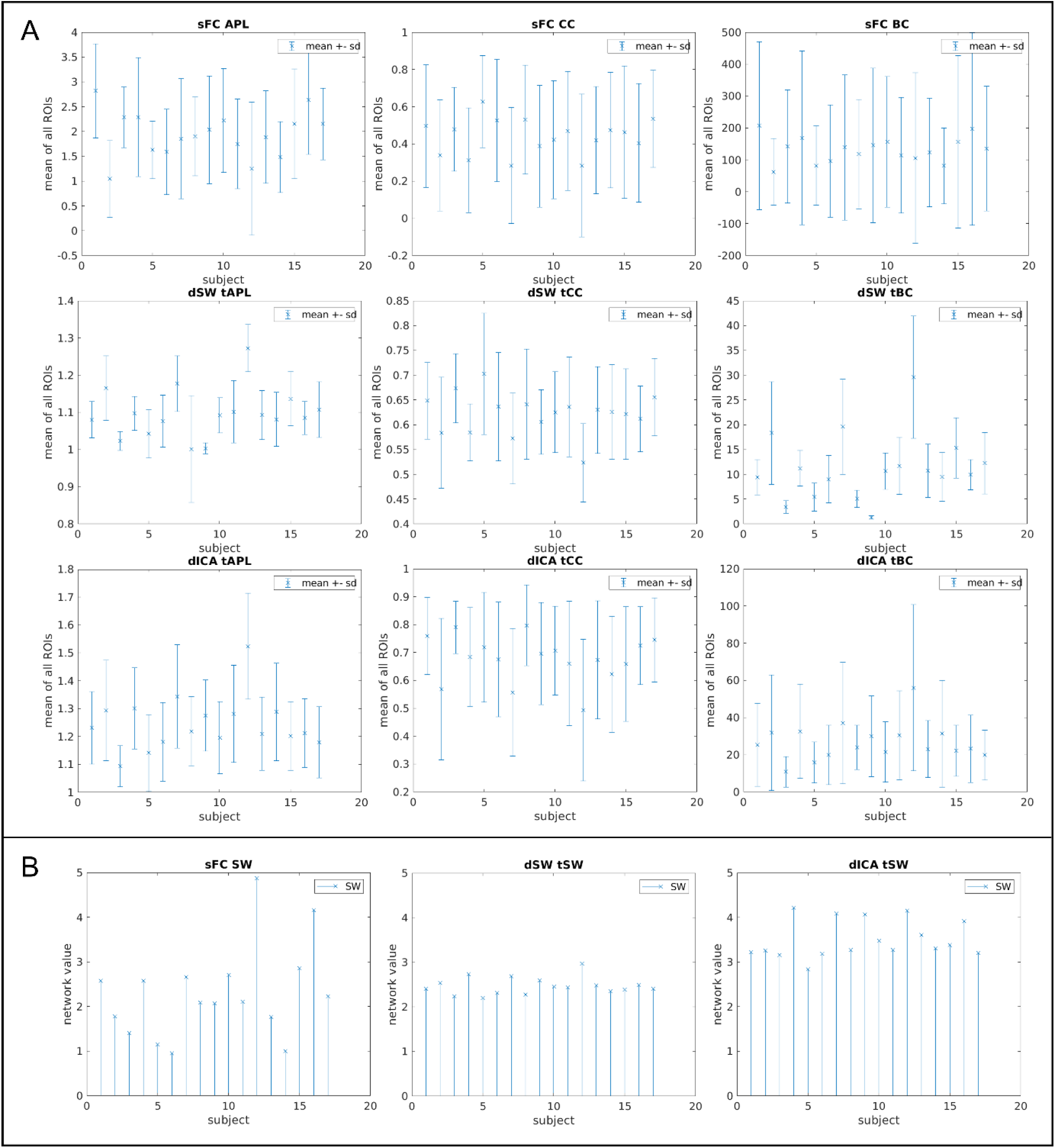
Distribution of network measures. (A) mean and standard deviation of the static, slow dynamic and fast dynamic average path length, clustering coefficient/temporal correlation coefficient and betweenness centrality of all subjects. (B) Stem plot of the static, slow dynamic and fast dynamic network small worldness.

### Predicting tic severity using LASSO regression

A predictor model for tic severity was created by applying linear regression with LASSO regularization to the network measures. This was preceded by a feature pre-selection step in which the Kendall’s tau between each measure of each ROI and the YGTSS was computed, and those with a p-uncorrected < 0.05 were selected for the regression model. Gender and medication were used as additional binary covariates for the model. The leave one out validated LASSO regression model yielded a minimum mean squared error (MSE) of 0.55 at a lambda (positive regularization parameter) of 0.019 (**Figure 4(A)**). **Figure 4(B)** shows the line between the predicted YGTSS and the actual YGTSS using this value of lambda. The coefficient of determination (R^2^), representing the goodness of fit, was found to be 0.97.

**Figure 4:**
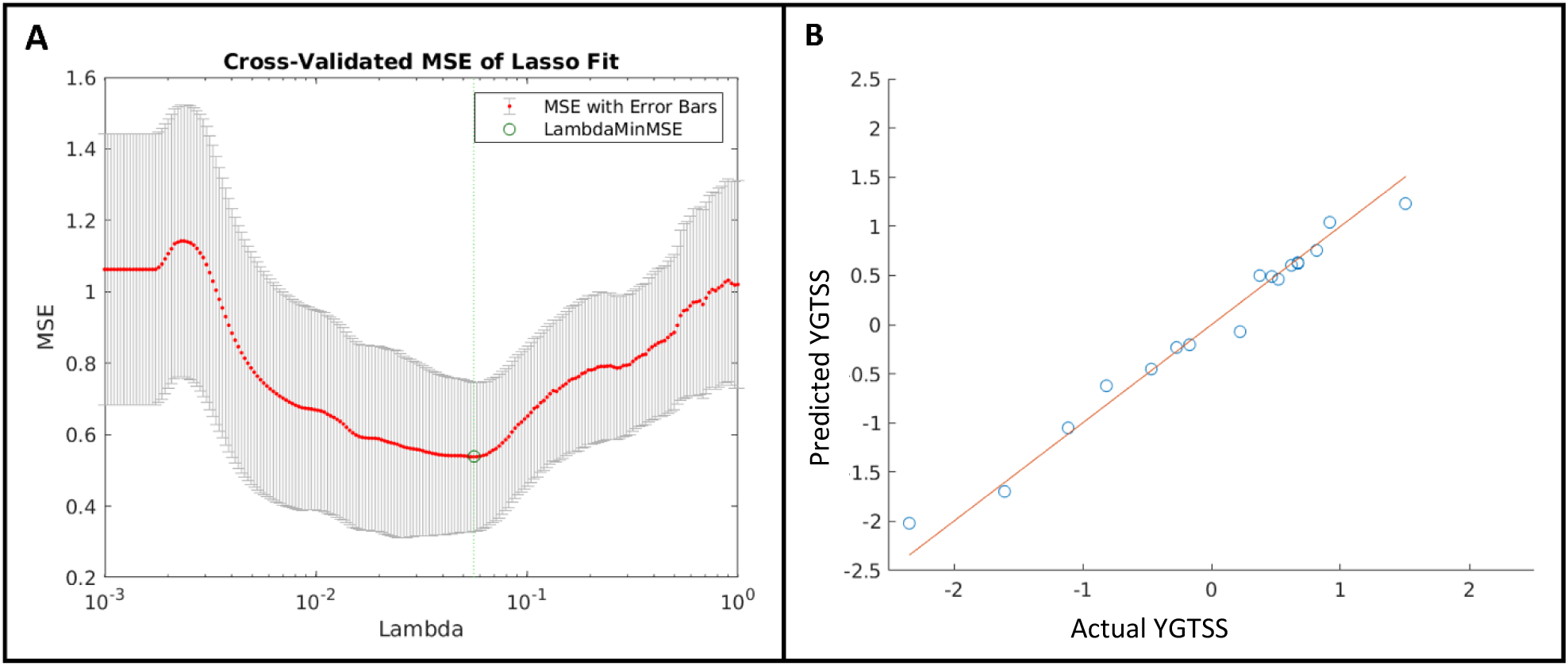
Results of prediction model. (A) mean squared error (MSE) with error bars for LOO-validation of LASSO regression for different values of lambda. The value of lambda with the minimum MSE (shown by the green line) was chosen as the optimal model, and the predicted YGTSS was plotted against the actual YGTSS as shown in (B). The coefficient of determination (R2), representing the goodness of fit, was found to be 0.97

### Feature space examination

Upon inspecting the feature space obtained by the model, eight predictors were identified, all at the region level, seven of which were dynamic network measures, and one was a static measure (Table 1). The clustering coefficient/ temporal correlation coefficient (CC/tCC) was the most relevant measure, with five out of eight identified features involving this measure. Two of the features involved the temporal betweenness centrality (tBC). Lastly, one of the identified features involved the average temporal path length (tAPL). The network-level features, the small worldness (SW) and the temporal small worldness (tSW), were not significantly correlated with the YGTSS during the feature selection step and thus were not included in the regression model. Both binary predictors, gender and medication, were reflected by the model in predicting tic severity; however they had very low weights (−0.15, 0.12 and 0.02, 0.001, respectively), indicating a small effect. The specific regions and the corresponding features are explained in further detail in the following subsections.

#### The static network (sFC network)

Only one of the predictors selected by the model belonged to the sFC network. This was the clustering coefficient of the right superior lateral occipital cortex (sLOC r). The Kendall’s correlation coefficient with the YGTSS was -0.37, and its weight in the model was -0.20. The static clustering coefficient is a measure of functional segregation, and a high value indicates a densely interconnected subnetwork organization engaged in specialized functioning. Thus, a negative correlation to the YGTSS sFC network indicates that a reduction in the functional segregation of the sLOC r may be associated with tics. The sLOC r was visualized on a 3D brain surface and can be seen in **Figure 5(A)**.

**Figure 5:**
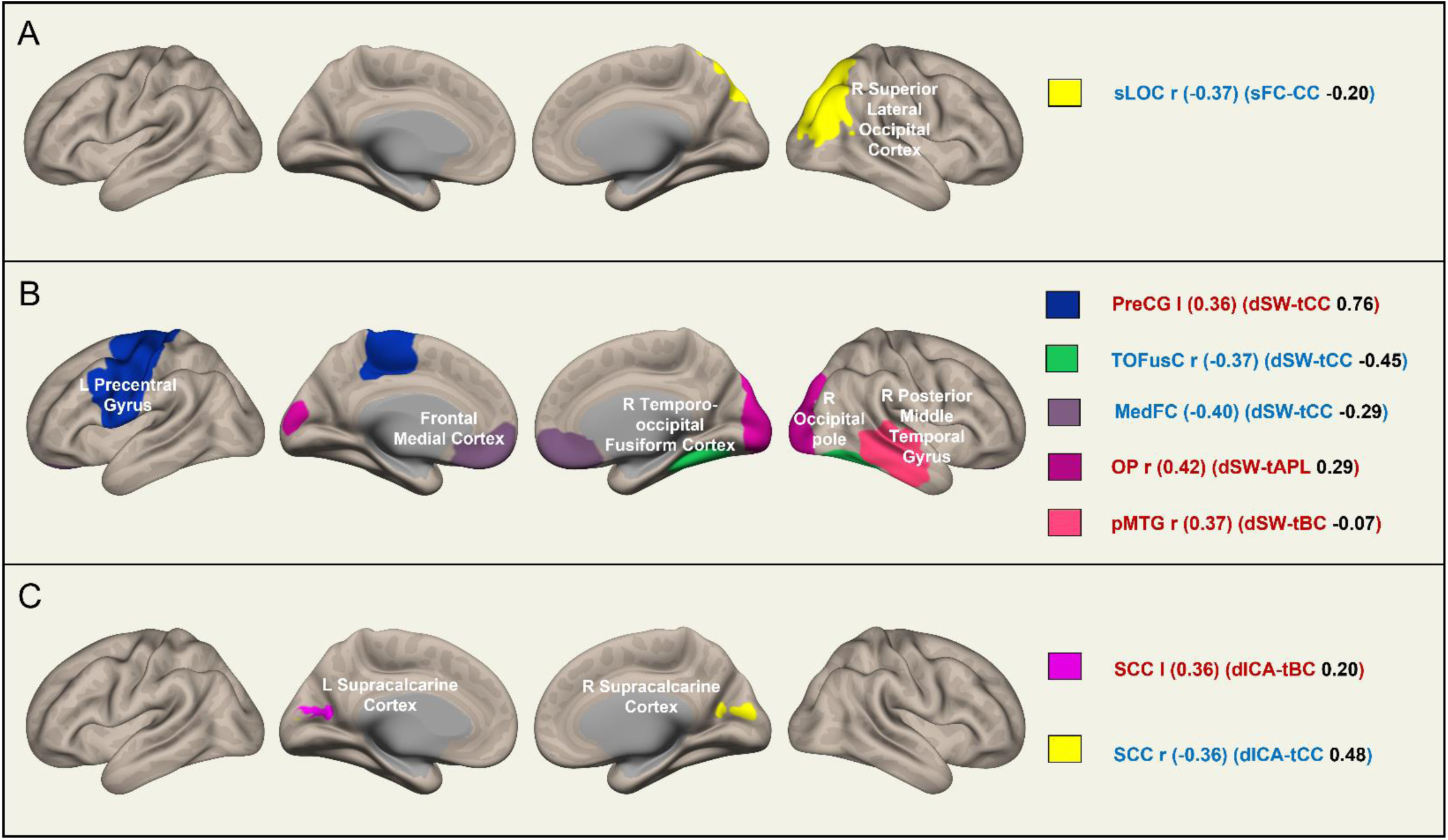
Predictors of tic severity visualized. The features obtained by the LASSO regression model visualized on the brain surface. Red indicates a positive correlation with YGTSS, and blue indicates a negative correlation. The value of the Kendall’s correlation coefficient is provided in the first parentheses, the network measure followed by its weight in the LASSO model is given in the second parentheses. The subfigures represent (A) static network, (B) slow dynamic network and (C) fast dynamic network. The effects of gender and medication were non-zero but had low weights of -0.15, 0.12 and 0.02, 0.001, respectively, indicating both had only a mild effect on the tic severity.

#### The direct slow dynamic network (dSW network)

Five of the eight identified predictors belonged to the dSW network. Of these, three were the temporal correlation coefficients (tCC) of the left precentral gyrus (PreCG l), the right temporooccipital fusiform cortex (TOFusC r) and the frontal medial cortex (MedFC), with Kendall’s correlation coefficients of 0.36, -0.37, -0.40 and model weights of 0.76, -0.45 and -0.29, respectively. As tics are typically motor in origin, it is no surprise that the primary motor cortex (i.e., PreCG l) appears as the predictor with the highest weight of 0.76 in the model. The temporal correlation coefficient indicates the stability of a node’s subnetwork during the dynamic course of information processing. This means that over a broad temporal scale, the precentral gyrus typically switches communication channels, receiving from and sending information to different subnetworks over the course of time. Failure to do so results in the constant subnetwork participation observed in relation to tic severity. In contrast, the negative correlation of the tCC of the TOFusC r and the MedFC to the YGTSS indicates that an unstable subnetwork organization of these two regions may be related to tics.

One predictor was the average temporal path length (tAPL) of the right occipital pole, with a Kendall’s correlation coefficient of 0.42 and model weight of 0.29. The last predictor of the dSW network was the temporal betweenness centrality (tBC) of the right posterior middle temporal gyrus (pMTG r), with a Kendall’s correlation coefficient of 0.37 and the least model weight of -0.07. The tAPL reflects how fast information is transferred between different subnetworks in time. A smaller temporal path length indicates that a node is well connected to other nodes across time and is thus active in dynamics, rapidly passing on information to other nodes. Consequently, our findings indicate selective connectedness and slow participation of the OP in time contributing to tic severity. Conversely, the tBC is the fraction of temporal short paths passing through a node, reflecting its importance in dynamic communication. Thus, a positive correlation indicates that greater involvement of the pMTG r in dynamic communication is instrumental in tic severity.

The predictors of the dSW network with their respective Kendall’s coefficients and weights were visualized on a 3D brain surface can be seen in **Figure 5(B)**.

#### The indirect fast dynamic network (dICA network)

Two of the obtained predictors were from the dICA network. One was the temporal correlation coefficient (tCC) of the right supracalcarine cortex (SCC r), with a Kendall’s tau of -0.36 and model weight of 0.48, and the was the temporal betweenness centrality (tBC) of the left supracalcarine cortex (SCC l), with a Kendall’s tau of 0.36 and model weight of 0.20. The temporal correlation coefficient (tCC) of the right supracalcarine cortex (SCC r) was found to be the second most important predictor obtained by the model after the dSW-tCC of the PreCG I. As explained above, the tCC reflects a node’s subnetwork stability, and the tBC is the reflection of a node’s importance. The dICA networks are an indirect measure of the dynamicity of communication at a fine temporal scale. Hence, our findings indicate that an unstable subnetwork organization, along with significant involvement of the SCC, contributes to tic severity. The predictors of the dSW network with their respective Kendall’s coefficients and weights were visualized on a 3D brain surface can be seen in **Figure 5(C)**.

## Discussion

This study investigates the functional network correlates of tic severity in Tourette syndrome (TS) by using machine learning. Resting-state fMRI data from TS patients were preprocessed and quality controlled, and static and dynamic functional connectivity were calculated. Network theory was applied to evaluate brain communication strategies, and the topological metrics were included in a LASSO regression model against the Yale global tic severity scale (the YGTSS) to identify the network features which yield the best predictability.

From the static network (sFC network), the clustering coefficient (CC) of the right superior lateral occipital cortex (sLOC-r) was identified as a predictor of tic severity. The lateral occipital cortex is known to be involved in attentional processing *(30)*. It has also been shown that attention to tics increases and away from them reduces their frequency *(31)*. Furthermore, Misirlisoy and colleagues *(31)* have proposed attentional modulation of motor noise as a contributing factor to tic generation, and Adolphs and Spezio have explained the role of the amygdala in controlling inward attention (towards the body and encoded emotional associations), proposing that malfunction in these networks leads to negative ideation and sensations accompanying mental illness *(32)*. The negative correlation of the CC to the YGTSS indicates a lack of adequate connections in the subnetwork of the sLOC-r in relation to tics, perhaps leading to the improper attentional modulation observed in tics.

Most of the identified predictors belonged to the slow dynamic network (dSW network). Of these, the temporal correlation coefficient (tCC) was the most relevant metric with three significant regions, namely the left precentral gyrus (PreCG-l), the frontal medial cortex (MedFC) and the right temporo-occipital fusiform cortex (TOFusC-r). In contrast, the tCC of the PreCG-l was positively correlated with the YGTSS, indicating a consistency in the subnetwork of the PreCG-l during dynamic communication contributing to tic severity. Conversely, the tCC of the MedFC and TOFuSC-r were negatively correlated with the YGTSS, indicating that changes in the subnetworks of these two regions during dynamic communication may be instrumental in tic severity. The preCG is the primary motor cortex that constantly communicates with other motor areas and subcortical brain regions to plan and execute movements *(33)*. Thus, the lack of network switching observed in dynamics indicates support for the lack of inhibition hypothesis *(8, 34)*. In normal brain functioning, spontaneous involuntary movements would be constantly balanced by inhibitory signals from other brain regions (observed as changing subnetworks in dynamics), and this failure of inhibitory control (observed as a constant subnetwork in dynamics) would lead to the sustained motor action observed in tics. Consistent activations in the pre-central gyrus have been reported 2 s and 1 s prior to tic onset and at tic onset *(20)*.

The frontal medial cortex (overlapping with the ventromedial prefrontal cortex (vmPFC)) is a key region of the default mode network and is involved in many critical networks and functions, such as self-referential processing *(35)*, personal and social decision making *(36)*, emotional regulation through the amygdala *(37, 38)* and processing of risk and fear *(39)*. Unlike the precentral gyrus, this finding, together with all the critical functions of the vmPFC, indicates the necessity of a stable subnetwork organization for healthy functioning, in which the vmPFC constantly receives and integrates inputs from all its different branches. The vmPFC has also been shown to be involved in exerting inhibitory control over the pre-SMA *(40)*. The pre-SMA is known to be involved in the cognitive control of actions that require inhibition or switching *(41)*. This complements the finding relating to the dSW-tCC in the preCG, wherein a dysfunction in one leads to a dysfunction in the other.

The TOFusC in our atlas corresponds to the subregion of the fusiform cortex specializing in face perception known as the fusiform face area (FFA) *(42)*. It has been proposed that the FFA encodes abstract semantic information associated with faces, which is then later retrieved for social computations *(43)*. The social dimension of tics has been emphasized in literature *(10)*, where it is proposed that TS is a disorder of the social decision-making network (SDM hypothesis). In this recent review article *(10)*, Albin has shed light on the possibility of tics being distorted social signals, emphasizing the role of typical tic movements, such as head, neck, facial and hand movements, in non-verbal emotional communication *(44)*. The framework of this new hypothesis puts together different pieces of the complex puzzle, explaining the cardinal features of tic disorders, such as the nature of tics, sex disparity and natural course of the illness, together with the basal ganglia and amygdala abnormalities observed in imaging *(9, 45, 46)* and post-mortem studies *(10)*. In this paper, Albin has further highlighted the three-fold role of the amygdala in social processing, i.e. in social perception, affliction and aversion, proposing the idea that the performance of the amygdala in the social functioning networks centred around it is altered by task engagement and attentional loading, thus explaining the modulation of tics during task engagement. The involvement of the amygdala in social processing has also been highlighted by Adolphs and Spezio *(32)*. They have put forth the idea that the amygdala attentionally modulates the visual and somatosensory cortices, directing visuospatial attention to face gaze, thus guiding contextual social behaviour. This framework brings together the somatosensory, visual and attentional networks under the umbrella of the amygdala, making it the key background integrator of all these different networks. Thus, our findings relating to sLOC and TOFusC indicate that the sLOC and the TOFusC could be working in tandem via the amygdala, causing attention to be directed to abstract semantic information stored in relation to facial expressions of present or past experiences and further motivating social behaviour. An inadequate subnetwork organization of one and the unstable subnetwork organization of the other could together lead to misinformed social signals.

The other two predictors of the dSW network were the average temporal path length (tAPL) of the right occipital pole (OP-r) and the temporal betweenness centrality (tBC) of the right posterior middle temporal gyrus (pMTG-r). These predictors suggest that delayed dynamic information transfer through the OP-r and increased involvement of the pMTG-r may be instrumental in increasing tic severity. A recent meta-analysis on the neuroimaging of the Cyberball game showed a reliable involvement of the OP in the experience of social exclusion *(47)*. Within the framework of the SDM hypothesis, this could indicate the lack of adequate connections to counter the experience of social exclusion, leading to a sustained social exclusion experience, which, once triggered, misinforms other networks to act on it. An H_2_^15^O-PET study has shown the MTG to be active in face perception and the posterior region has been specifically associated in the perception of fear *(48)*. In a previous study, we showed increased connectivity of the pMTG-r with several basal ganglia regions, such as the caudate, putamen and the pallidum in TS patients as compared to healthy controls *(45)*. Another study has shown that basal ganglia output modulated by striatal dopamine regulates social context-dependent behaviour *(49)*. In addition to this, Pourtois and colleagues *(48)* have shown the amygdala to be consistently activated in response to fearful faces. In one of our previous studies on TS using an emotional face task *(50)*, we showed amygdala hypersensitivity in TS patients as compared to healthy controls in response to all emotions. The activation was higher for negative emotions, i.e., angry and fearful, but neutral when compared to happy emotions. Taken together, these findings suggest that fear is overperceived by TS patients, and that this misperceived fear is quickly communicated to the basal ganglia by the pMTG, thus motivating misinformed social behaviour leading to tics. Consequently, the increased importance of the pMTG supports the suggestion that TS patients with more severe tics have higher levels of misperceived fear via the pMTG during dynamic communication.

Finally, the indirect fast dynamic network (dICA network) reflected the involvement of the supracalcarine cortex (SCC) in the severity of tics. Lower subnetwork stability (tCC of SCC-r) or higher node importance (tBC of SCC-l) was found to be related to higher severity of tics. The SCC, together with the occipital pole (OP), is the location of the primary visual cortex *(51)*; however, preferential activation to face-targets over non-face visual stimuli have been reported in the SCC in an fMRI study *(52)*. This indicates its role in social processing and is in line with the SDM hypothesis of TS *(10)*. An unstable subnetwork organization, while playing a key role in information transfer, could indicate misinterpreted social signals (such as facial expressions) being transmitted to other subnetworks, driving inappropriate social behaviour, leading to tics *(10)*.

In addition to the primary motor cortex and prefrontal cortex, our findings emphasize the role of several novel regions of the temporo-occipital cortex in TS. The temporo-occipital cortex is most well-known for its function as the visual cortex; however, more recently, its involvement in social processing *(32, 52)* and psychiatric disorders, such as major depression *(53)*, have come to light. These findings point to a new dimension of understanding that is in line with the emerging hypothesis of TS as a disorder of the social decision-making network *(10)*. A proposed visual social processing network mediated by the amygdala *(32)* involves communication between the regions obtained in our study through the amygdala. Thus, our network-based analysis approach helps in understanding how information is processed (both dynamically and overall) by each specific region engaged in this network in relation to tics in TS. It is interesting to note that, while several studies have shown a direct implication of the amygdala in TS *(9, 34)*, our study demonstrates that the network properties of the amygdala itself are not direct predictors of tic severity, rather, the regions it communicates with in the social processing context are. This broadens our understanding on the roles played by other regions in the network in addition to the amygdala itself.

## Limitations

The main limitation of this study is the small sample size. However, despite this, we observed convergence during leave one out validation, indicating the strength of our findings. Future studies with larger sample sizes could be used to replicate the findings and to explore network dynamics in Tourette syndrome in further detail.

## Data Availability

All data produced in the present study are available upon reasonable request to the authors.

## Acknowledgements

We thank the German Tourette’s Association for their support and travel funds and all participating Tourette’s patients and their families for their cooperation, Ms. Claire Rick for her editorial input on writing the manuscript and the Medical Technical Assistants at the Forschungszentrum, Jülich for assistance with data acquisition. This work is part of the Dr. rer. medic thesis of Ms. Shukti Ramkiran.

## Methods

### Participants

Following prior informed consent, a total of 36 adult patients fulfilling the DSM-IV-TR *(54)* criteria for TS participated in this study. Of these, five patients additionally suffered from obsessive-compulsive disorder (OCD), and two from attention-deficit-hyperactivity disorder (ADHD), as per the DSM-IV classification. Fourteen of the patients were on medication. Tic severity was measured using the YGTSS. Subsets of subjects from this dataset focusing on other analysis strategies have been previously published in *(4, 20, 46)*. Subjects with incomplete imaging or demographic data or with images corrupted by artefacts (motion, coverage, susceptibility etc.) were excluded from further analyses. After exclusion, 17 patients (12 male, 5 female, age: 32 ± 11 years) were subjected to further analyses, demographic details of which can be found in **Table 1**. All patients had normal or corrected-to-normal vision, no hearing loss and reported a strong right-hand preference *(46)*. The study was conducted according to the Declaration of Helsinki and under granted approval from the ethics committee of the medical faculty RWTH Aachen, Germany.

### Data acquisition

The image acquisition protocol comprised structural MRI, resting-state fMRI, task fMRI and diffusion MRI sessions acquired on a 1.5T whole-body MR system (Sonata, Siemens, Germany) at the Forschungszentrum Jülich. An MR-compatible video camera system was used to monitor tics in TS patients during the scanning sessions *(19)*. The resting-state fMRI data were acquired using a T2*-weighted echo-planar imaging sequence (scanning parameters: TE = 60 ms, TR = 3200 ms, flip angle = 90°, 30 axial slices 4 mm thick, FOV = 200 mm, in-plane resolution = 3.125 mm × 3.125 mm, 12 mins 220 volumes, eyes closed) and structural MRI acquired using a T1-weighted gradient-echo MP-RAGE sequence (scanning parameters: TI = 1200 ms, TR = 2200 ms, TE = 3.93 ms, 15° flip angle, FOV = 256 × 256 mm^2^, matrix size = 256 × 256, 176 sagittal slices generated, slice thickness = 1 mm, resolution = 1 mm isotropic) were used for further investigation in this study.

### Data pre-processing

The data were processed using standard pre-processing pipelines in CONN v20.b *(26)*, based on SPM12 *(55)*. Functional pre-processing involved the following steps: realignment and unwarp (for motion and field map correction), translation of the image centre (to the origin 0,0,0), slice-timing correction, outlier scan detection and scrubbing (using ART: artefact removal toolbox, parameters: global-signal z-value threshold: 5, subject-motion threshold: 0.9mm), and spatial normalization to an MNI152 (2 mm) template (using SPM’s unified segmentation *(56)*, parameters: functional target resolution 2mm) and functional smoothing (FWHM 8mm). Pre-processing of the structural images involved the following steps: translation of the image centre (to the origin 0,0,0), segmentation and normalization into MNI-space (using SPM’s unified segmentation *(56)*, parameters: structural target resolution: 1mm). Pre-processing was followed by nuisance regression of the following confounds: noise components of WM and CSF (aCompCor: first five principal components of time series *(26)*), estimated subject-motion parameters (six realignment parameters and their first derivatives), outlier scans identified (scrubbing) and the effect of rest (to compensate for initial magnetization transient). This was followed by temporal band-pass filtering at 0.008–0.09 Hz (to minimize the impact of physiological noise stemming from respiration and heartbeat) and linear detrending. (**Figure 6(A) outlines the pre-processing steps)**

**Figure 6:**
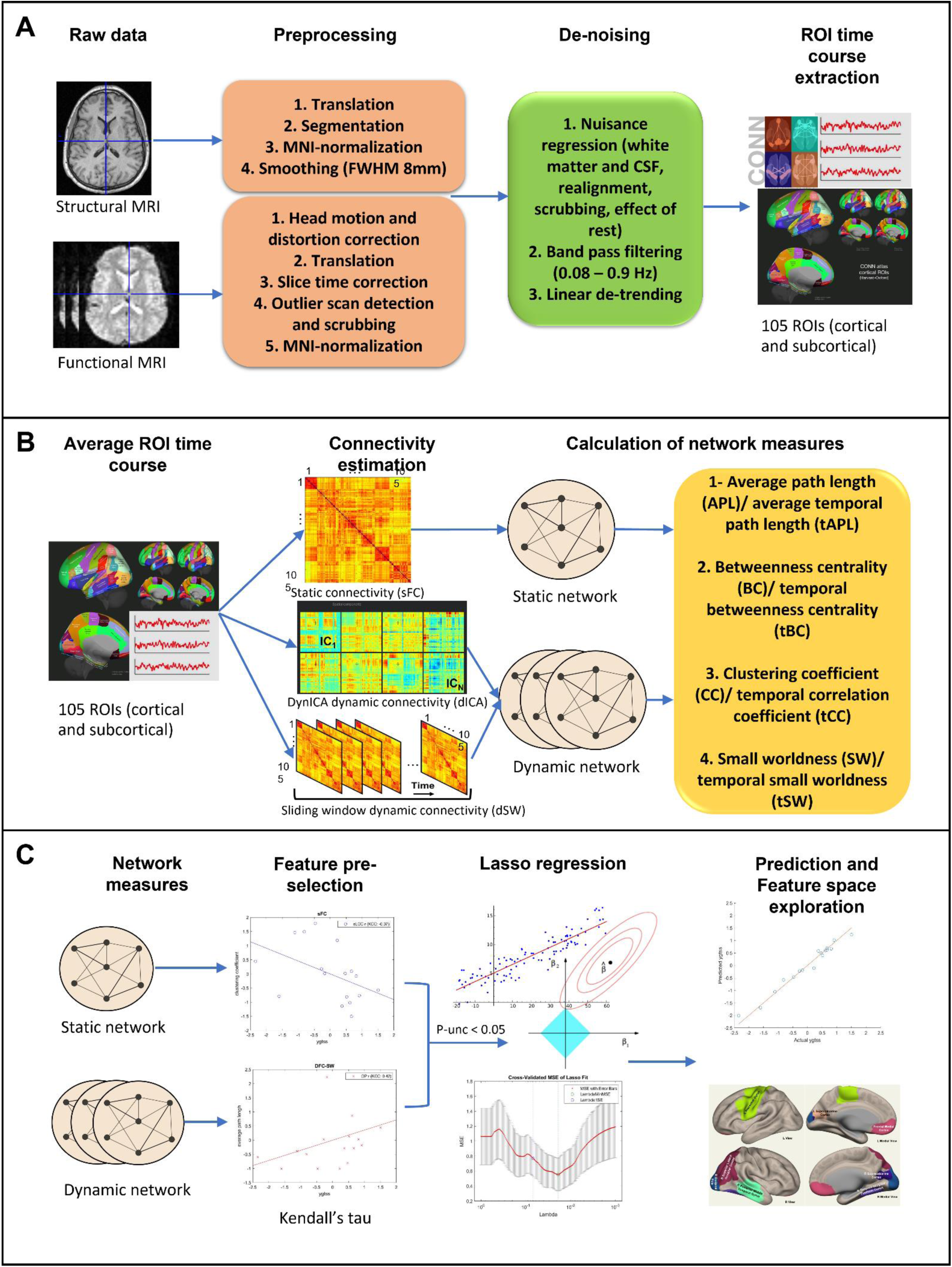
Data processing and analysis pipeline. (A) default preprocessing and denoising pipeline using CONN; (B) Static and dynamic network analysis: calculation of static functional connectivity(sFC), dynamic ICA based dynamic functional connectivity (dICA) and sliding window dynamic connectivity (dSW) followed by application of static and dynamic graph theory; (C) feature selection and prediction: feature pre-selection using p-unc<0.05 on Kendall’s tau between the YGTSS and each network measure, selected features are then fed to a leave one out (LOO-) LASSO regression model to obtain optimal parameters for the prediction of the YGTSS; finally the weights of the features in the optimal model are investigated.

### Static and dynamic network analyses

All connectivity analyses were performed using CONN v20.b. For static connectivity (sFC), an ROI-based functional connectivity model (bivariate correlation) was specified in the 1^st^-level analysis. The model included 105 regions covering the cortex (91 regions) and subcortex (14 regions) provided with CONN (parcellations as per the Harvard-Oxford cortical and subcortical maximum likelihood atlases). The cerebellum was excluded from the ROIs due to coverage artefacts in the images. Normalization of correlation values was performed using Fisher’s z-transformation.

For dynamic connectivity, two approaches were employed. The direct approach involved decomposing each session into 24 sliding windows of length 100 s and step size 25 s. Each window comprised 35 fMRI volumes. Following this, ROI-based functional connectivity was obtained for each sliding window in a similar way to the static connectivity approach (dSW).

The indirect dynamic connectivity, or dynamic ICA approach (dICA), is a relatively new approach that was introduced with the CONN toolbox. Here, the dynamic connectivity is estimated by first obtaining the different modulatory circuits and the rate of connectivity change between the ROIs at the group level, which are then back-projected to the individual level in a gPPI model *(18)*. The computation involved group-level ICA decomposition of the ROI-ROI connectivity timeseries into 20 components (selected by default in CONN), removal of ICs with spatial kurtosis < 3 (resulted in removal of five components), gPPI back-projection of the remaining ICs to the individual subject level followed by temporal smoothing of 10 s.

All the connectivity matrices were then imported into MATLAB R2021a, converted to raw correlation values and binarized using an absolute threshold of 0.45 (correlation values >0.45 or <-0.45 were retained). The value 0.45 was chosen after evaluating the network global efficiency distributions for optimal separation between random and ordered networks (maintaining optimal small-worldness) using CONN’s variable threshold explorer. Brain networks have been typically shown to be small world in nature *(57)*. Static and dynamic graph theory was applied to the binary adjacency matrices. Binary adjacency matrices can be visualized as binary graphs with ROIs as nodes and the connections between them as the edges of the graphs. An edge exists if the connectivity between them is greater than the applied threshold (in our case, correlation >0.45 or <-0.45). In this context, the following metrics were calculated:

1. **Average path length**: The path length is defined as the number of edges between two nodes (minimum number of edges for shortest path length), and the average path length (APL) of a given node is the average of the shortest path lengths between that node and all other nodes in the network. It reflects the functional integration ability and the speed of serial communication through a node *(22, 25)*. The temporal equivalent of the shortest path length is called the latency or the minimum number of time points that need to pass before the information can travel from one node to the other. Thus, temporal average path length (tAPL) is the average of the latencies between that node and all other nodes in the network. As before, it reflects the speed of communication in the dynamic network *(27)*.
2. **Clustering coefficient / temporal correlation coefficient**: Clustering coefficient (CC) is defined as the proportion of a node’s neighbours that are also neighbours of each other. It is a measure of functional segregation reflecting the extent of the specialized information processing a node is involved in. It reflects the average density and intra-connectedness of the node’s subnetwork *(58)*. Temporal correlation coefficient (tCC) is the average topological overlap of a node’s neighbours between two successive time points *(27)*. Based on this definition, tCC reflects the stability of a node’s subnetwork in dynamic performance. Nodes with a higher tCC would have a relatively stable subnetwork organization throughout the network dynamics.
3. **Betweenness centrality**: The fraction of shortest paths of the entire network passing through a given node is defined as its betweenness centrality (BC). It reflects a node’s importance and helps in identifying the hubs of a network *(24)*. Similarly, temporal betweenness centrality (tBC) is the fraction of the fastest paths passing through the node and reflects its importance in the dynamic network *(27)*.
4. **Small worldness:** Small worldness (SW) is a network property defined as the ratio of the clustering coefficient to the characteristic path length compared to random networks *(22)*. Small world networks combine high clustering of regular networks with short path lengths of random networks and lead to more efficient information processing. Similarly temporal small worldness (tSW) is defined as the ratio of the temporal correlation coefficient to the characteristic temporal path length when compared to null models *(27)*. (**Figure 6(B) outlines the network analyses)**

### Feature selection and prediction

After calculation of network and node metrics for all the ROIs, the Kendall’s coefficient of concordance with the YGTSS was calculated for each measure. Without correction for multiple comparisons, the measures that were significantly correlated (p-value < 0.05) were selected as features for the YGTSS prediction model. (**Supplementary Table S1 lists the Kendall’s tau for each measure)**. For this purpose, a leave-one-out (LOO) validated LASSO regression model was selected, in which the network features were used as continuous independent variables. The Fisher’s z transformed YGTSS was the continuous dependent variable, and gender and medication were categorical variables. LASSO regression applies a regularization term to linear regression in order to perform variable selection, thereby assigning weights to predictor variables in their order of significance and setting the weights of non-significant predictor variables to 0. Hereafter, the non-zero predictor variables identified by the model were inspected and have been discussed further. (**Figure 6(C) outlines the steps of the prediction model)**

## Data Availability

All data are available for research purposes only upon request from the corresponding author. The data cannot be made publicly available due to ethical concerns regarding patient data.

## Code Availability

All code is available upon request from the corresponding author.

## References

1. A. E. Cavanna, S. Servo, F. Monaco, M. M. Robertson, The Behavioral Spectrum of Gilles de la Tourette Syndrome, J. Neuropsychiatry Clin. Neurosci. 21, 13–23 (2009).

2. M. M. Robertson, V. Eapen, H. S. Singer, D. Martino, J. M. Scharf, P. Paschou, V. Roessner, D. W. Woods, M. Hariz, C. A. Mathews, R. Črnčec, J. F. Leckman, Gilles de la Tourette syndrome, Nat. Rev. Dis. Prim. 3, 16097 (2017).

3. D. S. Bassett, C. H. Xia, T. D. Satterthwaite, Understanding the Emergence of Neuropsychiatric Disorders with Network Neuroscience, Biol. psychiatry. Cogn. Neurosci. neuroimaging 3, 742 (2018).

4. S. Ramkiran, L. Heidemeyer, A. Gaebler, N. J. Shah, I. Neuner, Alterations in basal ganglia-cerebello-thalamo-cortical connectivity and whole brain functional network topology in Tourette’s syndrome, NeuroImage Clin. 24 (2019), doi:10.1016/j.nicl.2019.101998.

5. H. Wen, Y. Liu, I. Rekik, S. Wang, Z. Chen, J. Zhang, Y. Zhang, Y. Peng, H. He, Combining Disrupted and Discriminative Topological Properties of Functional Connectivity Networks as Neuroimaging Biomarkers for Accurate Diagnosis of Early Tourette Syndrome ChildrenMol. Neurobiol., 1–19 (2017).

6. Y. Worbe, C. Malherbe, A. Hartmann, M. Pélégrini-Issac, A. Messé, M. Vidailhet, S. Lehéricy, H. Benali, Functional immaturity of cortico-basal ganglia networks in Gilles de la Tourette syndrome, Brain 135, 1937–1946 (2012).

7. R. L. Albin, J. W. Mink, Recent advances in Tourette syndrome researchTrends Neurosci. 29, 175–182 (2006).

8. G. M. Jackson, A. Draper, K. Dyke, S. E. Pépés, S. R. Jackson, Inhibition, Disinhibition, and the Control of Action in Tourette Syndrome, Trends Cogn. Sci. 19, 655–665 (2015).

9. I. Neuner, T. Kellermann, T. Stöcker, T. Kircher, U. Habel, J. N. Shah, F. Schneider, Amygdala hypersensitivity in response to emotional faces in Tourette’s patients, World J. Biol. Psychiatry 11, 858–872 (2010).

10. R. L. Albin, Tourette syndrome: A disorder of the social decision-making network Brain 141, 332–347 (2018).

11. J. F. Leckman, M. A. Riddle, M. T. Hardin, S. I. Ort, K. L. Swartz, J. Stevenson, D. J. Cohen, The Yale Global Tic Severity Scale: Initial Testing of a Clinician-Rated Scale of Tic Severity, J. Am. Acad. Child Adolesc. Psychiatry 28, 566–573 (1989).

12. E. A. Storch, A. S. De Nadai, A. B. Lewin, J. F. McGuire, A. M. Jones, P. J. Mutch, R. D. Shytle, T. K. Murphy, Defining treatment response in pediatric tic disorders: a signal detection analysis of the Yale Global Tic Severity Scale, J. Child Adolesc. Psychopharmacol. 21, 621–627 (2011).

13. M. Haas, E. Jakubovski, C. Fremer, A. Dietrich, P. J. Hoekstra, B. Jäger, K. R. Müller-Vahl, Yale Global Tic Severity Scale (YGTSS): Psychometric Quality of the Gold Standard for Tic Assessment Based on the Large-Scale EMTICS Study, Front. Psychiatry 12, 98 (2021).

14. B. W. Mooneyham, M. D. Mrazek, A. J. Mrazek, K. L. Mrazek, D. T. Phillips, J. W. Schooler, States of mind: Characterizing the neural bases of focus and mind-wandering through dynamic functional connectivity, J. Cogn. Neurosci. 29, 495–506 (2017).

15. E. Damaraju, E. A. Allen, A. Belger, J. M. Ford, S. McEwen, D. H. Mathalon, B. A. Mueller, G. D. Pearlson, S. G. Potkin, A. Preda, J. A. Turner, J. G. Vaidya, T. G. Van Erp, V. D. Calhoun, Dynamic functional connectivity analysis reveals transient states of dysconnectivity in schizophrenia, NeuroImage Clin. 5, 298–308 (2014).

16. V. M. Vergara, A. R. Mayer, K. A. Kiehl, V. D. Calhoun, Dynamic functional network connectivity discriminates mild traumatic brain injury through machine learning, NeuroImage Clin. 19, 30–37 (2018).

17. R. M. Hutchison, T. Womelsdorf, E. A. Allen, P. A. Bandettini, V. D. Calhoun, M. Corbetta, S. Della Penna, J. H. Duyn, G. H. Glover, J. Gonzalez-Castillo, D. A. Handwerker, S. Keilholz, V. Kiviniemi, D. A. Leopold, F. de Pasquale, O. Sporns, M. Walter, C. Chang, Dynamic functional connectivity: Promise, issues, and interpretations, Neuroimage 80, 360–378 (2013).

18. Alfonso Nieto-Castanon, Handbook of functional connectivity Magnetic Resonance Imaging methods in CONN (2020).

19. I. Neuner, P. Wegener, T. Stoecker, T. Kircher, F. Schneider, N. J. Shah, Development and implementation of an MR-compatible whole body video system, Neurosci. Lett. 420, 122–127 (2007).

20. I. Neuner, C. J. Werner, J. Arrubla, T. StÃ¶cker, C. Ehlen, H. P. Wegener, F. Schneider, N. J. Shah, Imaging the where and when of tic generation and resting state networks in adult Tourette patients, Front. Hum. Neurosci. 8, 362 (2014).

21. H. Wen, Y. Liu, I. Rekik, S. Wang, J. Zhang, Y. Zhang, Y. Peng, H. He, Disrupted topological organization of structural networks revealed by probabilistic diffusion tractography in Tourette syndrome children, Hum. Brain Mapp. 38, 3988–4008 (2017).

22. O. Sporns, Structure and function of complex brain networks, Dialogues Clin. Neurosci. 15, 247–262 (2013).

23. E. Bullmore, O. Sporns, Complex brain networks: graph theoretical analysis of structural and functional systems., Nat. Rev. Neurosci. 10, 186–98 (2009).

24. M. Rubinov, O. Sporns, Complex network measures of brain connectivity: Uses and interpretations, Neuroimage 52, 1059–1069 (2010).

25. S. Achard, E. Bullmore, Efficiency and Cost of Economical Brain Functional Networks, PLoS Comput. Biol. 3, e17 (2007).

26. S. Whitfield-Gabrieli, A. Nieto-Castanon, Conn: A Functional Connectivity Toolbox for Correlated and Anticorrelated Brain Networks, Brain Connect. 2, 125–141 (2012).

27. A. E. Sizemore, D. S. Bassett, Dynamic graph metrics: Tutorial, toolbox, and taleNeuroimage 180, 417–427 (2018).

28. D. S. Bassett, E. Bullmore, Small-world brain networksNeuroscientist 12, 512–523 (2006).

29. A. E. Sizemore, D. S. Bassett, Dynamic graph metrics: Tutorial, toolbox, and tale Neuroimage180, 417–427 (2018).

30. S. O. Murray, E. Wojciulik, Attention increases neural selectivity in the human lateral occipital complex, Nat. Neurosci. 7, 70–74 (2004).

31. E. Misirlisoy, V. Brandt, C. Ganos, J. Tübing, A. Münchau, P. Haggard, The relation between attention and tic generation in Tourette syndrome., Neuropsychology 29, 658–65 (2015).

32. R. Adolphs, M. Spezio, Chapter 20 Role of the amygdala in processing visual social stimuliProg. Brain Res. 156, 363–378 (2006).

33. G. E. Alexander, M. R. Delong, P. L. Strick, Parallel Organization of Functionally Segregated Circuits Linking Basal Ganglia and Cortex, Annu. Rev. Neurosci. 9, 357–381 (1986).

34. A. Lerner, A. Bagic, J. M. Simmons, Z. Mari, O. Bonne, B. Xu, D. Kazuba, P. Herscovitch, R. E. Carson, D. L. Murphy, W. C. Drevets, M. Hallett, Widespread abnormality of the γ-aminobutyric acid-ergic system in Tourette syndrome, Brain 135, 1926–1936 (2012).

35. Y. I. Sheline, D. M. Barch, J. L. Price, M. M. Rundle, S. N. Vaishnavi, A. Z. Snyder, M. A. Mintun, S. Wang, R. S. Coalson, M. E. Raichle, The default mode network and self-referential processes in depression, Proc. Natl. Acad. Sci. U. S. A. 106, 1942–1947 (2009).

36. A. Bechara, D. Tranel, H. Damasio, Characterization of the decision-making deficit of patients with ventromedial prefrontal cortex lesions, Brain 123, 2189–2202 (2000).

37. J. Decety, K. J. Michalska, Neurodevelopmental changes in the circuits underlying empathy and sympathy from childhood to adulthood, Dev. Sci. 13, 886–899 (2010).

38. J. C. Motzkin, C. L. Philippi, R. C. Wolf, M. K. Baskaya, M. Koenigs, Ventromedial prefrontal cortex is critical for the regulation of amygdala activity in humans, Biol. Psychiatry 77, 276–284 (2015).

39. J. Hiser, M. Koenigs, The multifaceted role of ventromedial prefrontal cortex in emotion, decision-making, social cognition, and psychopathology, Biol. Psychiatry 83, 638 (2018).

40. J. Yu, P. Tseng, D. L. Hung, S. W. Wu, C. H. Juan, Brain stimulation improves cognitive control by modulating medial-frontal activity and preSMA-vmPFC functional connectivity, Hum. Brain Mapp. 36, 4004 (2015).

41. I. Obeso, N. Robles, E. Muñoz-Marrón, D. Redolar-Ripoll, Dissociating the role of the pre-SMA in response inhibition and switching: A combined online and offline TMS approach, Front. Hum. Neurosci. 0, 150 (2013).

42. N. Kanwisher, G. Yovel, The fusiform face area: a cortical region specialized for the perception of faces, Philos. Trans. R. Soc. B Biol. Sci. 361, 2109–2128 (2006).

43. R. T. Schultz, D. J. Grelotti, A. Klin, J. Kleinman, C. Van Der Gaag, R. Marois, P. Skudlarski, The role of the fusiform face area in social cognition: Implications for the pathobiology of autism, Philos. Trans. R. Soc. B Biol. Sci. 358, 415–427 (2003).

44. P. Ekman, W. V Friesen, P. Ellsworth, Emotion in the human face: Guidelines for research and an integration of findings (2013).

45. S. Ramkiran, L. Heidemeyer, A. Gaebler, N. J. Shah, I. Neuner, Alterations in basal ganglia-cerebello-thalamo-cortical connectivity and whole brain functional network topology in Tourette’s syndrome, NeuroImage Clin. 24 (2019), doi:10.1016/j.nicl.2019.101998.

46. C. J. Werner, T. Stöcker, T. Kellermann, H. P. Wegener, F. Schneider, N. J. Shah, I. Neuner, in European Archives of Psychiatry and Clinical Neuroscience, (2010), vol. 260.

47. L. Mwilambwe-Tshilobo, R. N. Spreng, Social exclusion reliably engages the default network: A meta-analysis of Cyberball, Neuroimage 227, 117666 (2021).

48. G. Pourtois, B. de Gelder, A. Bol, M. Crommelinck, Perception of facial expressions and voices and of their combination in the human brain, Cortex 41, 49–59 (2005).

49. A. Leblois, B. J. Wendel, D. J. Perkel, Striatal Dopamine Modulates Basal Ganglia Output and Regulates Social Context-Dependent Behavioral Variability through D1 Receptors, J. Neurosci. 30, 5730–5743 (2010).

50. I. Neuner, T. Kellermann, T. Stöcker, T. Kircher, U. Habel, J. N. Shah, F. Schneider, Amygdala hypersensitivity in response to emotional faces in Tourette’s patients, World J. Biol. Psychiatry 11, 858–872 (2010).

51. G. Leuba, R. Kraftsik, Changes in volume, surface estimate, three-dimensional shape and total number of neurons of the human primary visual cortex from midgestation until old age, Anat. Embryol. (Berl). 190, 351–366 (1994).

52. G. S. Dichter, J. N. Felder, J. W. Bodfish, L. Sikich, A. Belger, Mapping social target detection with functional magnetic resonance imaging, Soc. Cogn. Affect. Neurosci. 4, 59–69 (2009).

53. D.-Y. Liu, X. Ju, Y. Gao, J.-F. Han, Z. Li, X.-W. Hu, Z.-L. Tan, G. Northoff, X. M. Song, From Molecular to Behavior: Higher Order Occipital Cortex in Major Depressive Disorder, Cereb. Cortex (2021), doi:10.1093/CERCOR/BHAB343.

54. American Psychiatric Association, Diagnostic and statistical manual of mental disorders (2000; http://bvbr.bib-bvb.de:8991/F?func=service&doc_library=BVB01&local_base=BVB01&doc_number=025719079&line_number=0001&func_code=DB_RECORDS&service_type=MEDIA).

55. K. J. Friston, A. P. Holmes, K. J. Worsley, J. -P Poline, C. D. Frith, R. S. J. Frackowiak, Statistical parametric maps in functional imaging: A general linear approach, Hum. Brain Mapp. 2, 189–210 (1994).

56. J. Ashburner, K. J. Friston, Unified segmentation, Neuroimage 26, 839–851 (2005).

57. E. Bullmore, O. Sporns, The economy of brain network organization, Nat. Rev. Neurosci. 13, 336–349 (2012).

58. V. Latora, M. Marchiori, Efficient behavior of small-world networks., Phys. Rev. Lett. 87, 198701 (2001).

