## Supplementary Table S1 for "How brain networks tic: predicting tic severity through rs-fMRI dynamics in Tourette syndrome"

**Dynamic ICA – estimated dynamic functional connectivity (dICA)**

| Region | temporal correlation coefficien: Kendall's tau | p-value | average temporal path length: Kendall's tau | p-value | temporal betweenness centralit: Kendall's tau | p-value |
|---|---|---|---|---|---|---|
| Frontal Pole Right | -0.028203561 | 0.914432551 | 0.098852404 | 0.705831503 | 0.137339082 | 0.599142984 |
| Frontal Pole Left | 0.180257545 | 0.488739974 | -0.164544802 | 0.527985468 | 0.175352577 | 0.500837555 |
| Insular Cortex Right | -0.098099344 | 0.707979696 | -0.129679093 | 0.619853851 | -0.012262418 | 0.962744755 |
| Insular Cortex Left | -0.107909278 | 0.680164132 | -0.004178863 | 0.985659339 | 0.08828941 | 0.736148838 |
| Superior Frontal Gyrus Right | -0.251379569 | 0.330420538 | -0.038805362 | 0.882448341 | 0.03433477 | 0.89591734 |
| Superior Frontal Gyrus Left | -0.030656045 | 0.907020959 | -0.161109267 | 0.536753301 | 0.174126336 | 0.503883982 |
| Middle Frontal Gyrus Right | -0.240343393 | 0.352784473 | -0.094772591 | 0.717494403 | 0.06131209 | 0.815171399 |
| Middle Frontal Gyrus Left | -0.161863918 | 0.53482175 | 0.057265601 | 0.827190913 | -0.090741893 | 0.729075143 |
| Inferior Frontal Gyrus, pars triangularis Right | -0.291845548 | 0.255697691 | 0.081181057 | 0.756763152 | -0.152053983 | 0.56017219 |
| Inferior Frontal Gyrus, pars triangularis Left | -0.500306654 | 0.040825499 | 0.008044856 | 0.975553866 | 0.060085848 | 0.818809784 |
| Inferior Frontal Gyrus, pars opercularis Right | -0.133660356 | 0.609054423 | -0.159040622 | 0.542064111 | 0.166768885 | 0.522344507 |
| IFG oper l (Inferior Frontal Gyrus, pars opercularis L | -0.534641425 | 0.027027939 | -0.206255552 | 0.427059392 | 0.317596626 | 0.214157487 |
| PreCG r (Precentral Gyrus Right) | -0.228080975 | 0.378610298 | 0.04948279 | 0.850408764 | 0.150827741 | 0.563377227 |
| PreCG l (Precentral Gyrus Left) | 0.073574508 | 0.778994981 | -0.006452613 | 0.98039128 | 0.269773196 | 0.295028193 |
| TP r (Temporal Pole Right) | -0.121397938 | 0.64255182 | 0.370992441 | 0.142634686 | -0.035561012 | 0.892220091 |
| TP l (Temporal Pole Left) | -0.038013496 | 0.884831959 | 0.205320898 | 0.429203523 | -0.025751078 | 0.921850815 |
| aSTG r (Superior Temporal Gyrus, anterior division | -0.085836926 | 0.743242629 | 0.03582541 | 0.891423176 | -0.024524836 | 0.925562269 |
| aSTG l (Superior Temporal Gyrus, anterior division | -0.344573946 | 0.175603638 | -0.066801715 | 0.798927824 | -0.127529147 | 0.625716593 |
| pSTG r (Superior Temporal Gyrus, posterior divisior | 0.012262418 | 0.962744755 | -0.075214181 | 0.77418833 | 0.217044799 | 0.402717435 |
| pSTG l (Superior Temporal Gyrus, posterior division | -0.003678725 | 0.988820113 | -0.236424257 | 0.360927503 | 0.250153327 | 0.33286389 |
| aMTG r (Middle Temporal Gyrus, anterior division R | 0.01348866 | 0.959022034 | 0.061500801 | 0.814611788 | 0.376456233 | 0.13638905 |
| aMTG l (Middle Temporal Gyrus, anterior division L | -0.202329897 | 0.436102506 | -0.041410286 | 0.874614016 | -0.112814246 | 0.666398678 |
| pMTG r (Middle Temporal Gyrus, posterior division | -0.091968135 | 0.725545991 | -0.223434574 | 0.388661015 | 0.091968135 | 0.725545991 |
| pMTG l (Middle Temporal Gyrus, posterior division | -0.236664667 | 0.360429472 | 0.023646831 | 0.928220614 | -0.107909278 | 0.680164132 |
| toMTG r (Middle Temporal Gyrus, temporooccipita | -0.031882287 | 0.903317846 | 0.156855879 | 0.547698333 | 0.350705155 | 0.167536807 |
| toMTG l (Middle Temporal Gyrus, temporooccipita | 0.024524836 | 0.925562269 | -0.046169406 | 0.860329817 | -0.100551828 | 0.700991427 |
| aITG r (Inferior Temporal Gyrus, anterior division Ri | -0.212139831 | 0.41368987 | 0.070154178 | 0.789045801 | -0.117719213 | 0.652733157 |
| aITG l (Inferior Temporal Gyrus, anterior division Le | -0.362967573 | 0.152159806 | 0.115692855 | 0.658366352 | -0.259963262 | 0.313609781 |
| pITG r (Inferior Temporal Gyrus, posterior division R | -0.137339082 | 0.599142984 | 0.020628626 | 0.93736418 | 0.049049672 | 0.851704448 |
| pITG l (Inferior Temporal Gyrus, posterior division L | -0.138565323 | 0.595853798 | 0.017957431 | 0.945462703 | 0.008583693 | 0.973919696 |
| toITG r (Inferior Temporal Gyrus, temporooccipital | -0.278356889 | 0.279324367 | -0.060481762 | 0.817634682 | -0.133660356 | 0.609054423 |
| toITG l (Inferior Temporal Gyrus, temporooccipital | -0.169221368 | 0.516156614 | 0.18926601 | 0.466895444 | 0.003678725 | 0.988820113 |
| PostCG r (Postcentral Gyrus Right) | -0.028203561 | 0.914432551 | 0.070810717 | 0.787114035 | 0.101778069 | 0.697505734 |
| PostCG l (Postcentral Gyrus Left) | -0.141017807 | 0.589297674 | -0.012982169 | 0.960559587 | 0.257510778 | 0.318360465 |
| SPL r (Superior Parietal Lobule Right) | -0.273451921 | 0.288234406 | -0.184159853 | 0.479217271 | -0.02697732 | 0.918140885 |
| SPL l (Superior Parietal Lobule Left) | -0.147149016 | 0.573039516 | -0.242685193 | 0.347968917 | -0.03433477 | 0.89591734 |
| aSMG r (Supramarginal Gyrus, anterior division Rig | -0.202329897 | 0.436102506 | 0.060202584 | 0.818463265 | -0.098099344 | 0.707979696 |
| aSMG l (Supramarginal Gyrus, anterior division Left | 0.283261856 | 0.270584117 | -0.253843006 | 0.325543541 | 0.04782343 | 0.855374717 |
| pSMG r (Supramarginal Gyrus, posterior division Ri | -0.121397938 | 0.64255182 | -0.009000043 | 0.97265228 | 0.133660356 | 0.609054423 |
| pSMG l (Supramarginal Gyrus, posterior division Le | 0.203556139 | 0.433267183 | 0.077512012 | 0.767465374 | 0.006131209 | 0.981367826 |
| AG r (Angular Gyrus Right) | -0.230533458 | 0.373363679 | -0.074785087 | 0.775445464 | 0.147149016 | 0.573039516 |
| AG l (Angular Gyrus Left) | -0.011036176 | 0.966468204 | 0.028176891 | 0.914513187 | -0.069895783 | 0.78980641 |
| sLOC r (Lateral Occipital Cortex, superior division Ri | -0.361741331 | 0.153652534 | -0.274968452 | 0.285461459 | -0.055180881 | 0.833397631 |
| sLOC l (Lateral Occipital Cortex, superior division Le | -0.161863918 | 0.53482175 | 0.12058386 | 0.644799784 | -0.007357451 | 0.977642192 |
| iLOC r (Lateral Occipital Cortex, inferior division Rig | -0.305334208 | 0.233356236 | 0.094262536 | 0.718956705 | -0.080931959 | 0.757488449 |
| iLOC l (Lateral Occipital Cortex, inferior division Lef | -0.164316401 | 0.528566334 | 0.123673177 | 0.636284651 | 0.198651172 | 0.444665657 |
| ICC r (Intracalcarine Cortex Right) | -0.507664105 | 0.037496416 | -0.061787701 | 0.813761168 | 0.103004311 | 0.694025752 |
| ICC l (Intracalcarine Cortex Left) | -0.447578257 | 0.071620089 | -0.101957437 | 0.696996344 | 0.036787254 | 0.888524941 |
| MedFC (Frontal Medial Cortex) | -0.376456233 | 0.13638905 | 0.157056237 | 0.547180552 | -0.012262418 | 0.962744755 |
| SMA r (Juxtapositional Lobule Cortex -formerly Sup | -0.085836926 | 0.743242629 | -0.074019469 | 0.777689836 | 0.110361762 | 0.673269147 |
| SMA L(Juxtapositional Lobule Cortex -formerly Supp | -0.150827741 | 0.563377227 | 0.095183024 | 0.716318383 | 0.127529147 | 0.625716593 |
| SubCalC (Subcallosal Cortex) | -0.050275914 | 0.848037054 | 0.200520524 | 0.440303584 | -0.07480075 | 0.775399567 |
| PaCiG r (Paracingulate Gyrus Right) | -0.282035614 | 0.27275325 | -0.066627294 | 0.799442766 | 0.106683037 | 0.68362067 |
| PaCiG l (Paracingulate Gyrus Left) | 0.091968135 | 0.725545991 | -0.068996615 | 0.792454583 | 0.208461106 | 0.42202201 |
| AC (Cingulate Gyrus, anterior division) | 0.062538332 | 0.811536581 | -0.469967761 | 0.056965546 | 0.311465417 | 0.223624917 |
| PC (Cingulate Gyrus, posterior division) | 0.024524836 | 0.925562269 | -0.210368307 | 0.417691345 | 0.17044761 | 0.5130755 |
| Precuneous (Precuneous Cortex) | -0.196198688 | 0.450421764 | 0.193847071 | 0.455976417 | 0.019619869 | 0.940421887 |
| Cuneal r (Cuneal Cortex Right) | -0.615573374 | 0.008523584 | 0.051226393 | 0.845196422 | -0.360515089 | 0.155155198 |
| Cuneal l (Cuneal Cortex Left) | -0.171673852 | 0.510002995 | 0.04110048 | 0.875545196 | -0.234212184 | 0.365569868 |
| FOrb r (Frontal Orbital Cortex Right) | 0.014714902 | 0.955300113 | 0.317152067 | 0.214835096 | -0.038013496 | 0.884831959 |
| FOrb l (Frontal Orbital Cortex Left) | -0.212139831 | 0.41368987 | -0.220340884 | 0.395432857 | 0.022072352 | 0.932989463 |
| aPaHC r (Parahippocampal Gyrus, anterior division | -0.008583693 | 0.973919696 | 0.076402436 | 0.770709828 | -0.025751078 | 0.921850815 |
| aPaHC l (Parahippocampal Gyrus, anterior division | -0.295524274 | 0.249477184 | 0.205793658 | 0.428118295 | -0.294298032 | 0.251540063 |
| pPaHC r (Parahippocampal Gyrus, posterior divisior | 0.08828941 | 0.736148838 | 0.084780409 | 0.746304662 | 0.04782343 | 0.855374717 |
| pPaHC l (Parahippocampal Gyrus, posterior division | 0.051502156 | 0.844372606 | -0.177781165 | 0.494829999 | -0.022072352 | 0.932989463 |
| LG r (Lingual Gyrus Right) | -0.421827179 | 0.091692562 | 0.456150133 | 0.065715899 | -0.221949766 | 0.39190321 |

**Sliding window dynamic functional connectivity (dSW)**

| Region | temporal correlation coefficie: Kendall's tau | p-value | average temporal path length (t: Kendall's tau | p-value | temporal betweenness centrality: Kendall's tau | p-value |
|---|---|---|---|---|---|---|
| Frontal Pole Right | 0.04782343 | 0.855374717 | 0.313522876 | 0.220418546 | 0.360515089 | 0.155155198 |
| Frontal Pole Left | 0.057633365 | 0.826096984 | 0.202372859 | 0.436003008 | 0.289393065 | 0.259897822 |
| Insular Cortex Right | 0.138565323 | 0.595853798 | 0.391122623 | 0.120564771 | 0.326180319 | 0.20134434 |
| Insular Cortex Left | 0.350705155 | 0.167536807 | 0.465839621 | 0.059483157 | 0.207234864 | 0.424818826 |
| Superior Frontal Gyrus Right | 0.029429803 | 0.910725885 | 0.333785716 | 0.19041894 | 0.351931397 | 0.165953884 |
| Superior Frontal Gyrus Left | -0.031882287 | 0.903317846 | 0.315803095 | 0.216899697 | 0.228080975 | 0.378610298 |
| Middle Frontal Gyrus Right | 0.033108529 | 0.899616615 | 0.482675514 | 0.049712382 | 0.225628491 | 0.383897317 |
| Middle Frontal Gyrus Left | -0.023298594 | 0.929275176 | 0.32157714 | 0.208151806 | 0.141017807 | 0.589297674 |
| Inferior Frontal Gyrus, pars triangularis Right | -0.186388754 | 0.473819188 | 0.107744586 | 0.680628021 | 0.285714339 | 0.26627772 |
| Inferior Frontal Gyrus, pars triangularis Left | 0.120171696 | 0.645939027 | 0.338502802 | 0.183843045 | 0.217044799 | 0.402717435 |
| Inferior Frontal Gyrus, pars opercularis Right | 0.202329897 | 0.436102506 | 0.21886826 | 0.398678597 | 0.436542081 | 0.079782919 |
| IFG oper l (Inferior Frontal Gyrus, pars opercularis L | 0.190067479 | 0.464975812 | 0.411587316 | 0.100701418 | 0.217044799 | 0.402717435 |
| PreCG r (Precentral Gyrus Right) | 0.21827104 | 0.399998983 | 0.257189367 | 0.318986179 | 0.239117151 | 0.355321036 |
| PreCG l (Precentral Gyrus Left) | 0.450030741 | 0.069892611 | 0.126017455 | 0.629851625 | 0.496627929 | 0.042571778 |
| TP r (Temporal Pole Right) | -0.023298594 | 0.929275176 | 0.545661787 | 0.023469586 | 0.283261856 | 0.270584117 |
| TP l (Temporal Pole Left) | 0.12262418 | 0.639171259 | 0.114817002 | 0.6608066 | 0.239117151 | 0.355321036 |
| aSTG r (Superior Temporal Gyrus, anterior division | -0.039239738 | 0.881141218 | 0.127019872 | 0.627108477 | 0.472103093 | 0.055694842 |
| aSTG l (Superior Temporal Gyrus, anterior division | -0.364193815 | 0.150676993 | 0.304856081 | 0.234126226 | 0.17044761 | 0.5130755 |
| pSTG r (Superior Temporal Gyrus, posterior divisior | -0.020846111 | 0.936705057 | 0.206329841 | 0.42688921 | 0.337216495 | 0.185621085 |
| pSTG l (Superior Temporal Gyrus, posterior division | -0.220723524 | 0.394591846 | 0.375386392 | 0.137596825 | 0.329859044 | 0.196009784 |
| aMTG r (Middle Temporal Gyrus, anterior division R | -0.29307179 | 0.253613565 | 0.275078225 | 0.28526137 | 0.312691659 | 0.221710351 |
| aMTG l (Middle Temporal Gyrus, anterior division L | -0.451256982 | 0.069040442 | 0.049723766 | 0.849688034 | 0.337216495 | 0.185621085 |
| pMTG r (Middle Temporal Gyrus, posterior division | -0.141017807 | 0.589297674 | -0.064560667 | 0.805549965 | 0.464745642 | 0.060163967 |
| pMTG l (Middle Temporal Gyrus, posterior division | 0.069895783 | 0.78980641 | 0.21059257 | 0.417183658 | 0.289393065 | 0.259897822 |
| toMTG r (Middle Temporal Gyrus, temporooccipita | 0.024524836 | 0.925562269 | 0.045708282 | 0.86171213 | 0.556713777 | 0.020277394 |
| toMTG l (Middle Temporal Gyrus, temporooccipita | -0.117719213 | 0.652733157 | 0.513312817 | 0.035083992 | 0.326180319 | 0.20134434 |
| aITG r (Inferior Temporal Gyrus, anterior division Ri | -0.040465979 | 0.877452788 | 0.2516882 | 0.329807217 | 0.297976757 | 0.245383286 |
| aITG l (Inferior Temporal Gyrus, anterior division Le | -0.312691659 | 0.221710351 | 0.355783382 | 0.161047087 | 0.282035614 | 0.27275325 |
| pITG r (Inferior Temporal Gyrus, posterior division R | -0.116492971 | 0.656139965 | 0.302529 | 0.237896821 | 0.283261856 | 0.270584117 |
| pITG l (Inferior Temporal Gyrus, posterior division L | -0.256284536 | 0.320751547 | 0.472909632 | 0.055220423 | 0.202329897 | 0.436102506 |
| toITG r (Inferior Temporal Gyrus, temporooccipital | -0.208461106 | 0.42202201 | 0.355433288 | 0.161488947 | 0.152053983 | 0.56017219 |
| toITG l (Inferior Temporal Gyrus, temporooccipital | -0.175352577 | 0.500837555 | 0.369142835 | 0.144792759 | 0.24524836 | 0.342741321 |
| PostCG r (Postcentral Gyrus Right) | 0.02697732 | 0.918140885 | 0.55549381 | 0.020612116 | 0.213366073 | 0.410932019 |
| PostCG l (Postcentral Gyrus Left) | 0.123850422 | 0.6357974 | 0.206562025 | 0.426357551 | 0.262415745 | 0.308901139 |
| SPL r (Superior Parietal Lobule Right) | 0.071122024 | 0.786198469 | 0.225537724 | 0.384093764 | 0.286940581 | 0.264140461 |
| SPL l (Superior Parietal Lobule Left) | 0.045370947 | 0.862723602 | 0.21084063 | 0.416622482 | 0.073574508 | 0.778994981 |
| aSMG r (Supramarginal Gyrus, anterior division Rig | 0.161863918 | 0.53482175 | -0.010553186 | 0.967934975 | 0.375229991 | 0.137774008 |
| aSMG l (Supramarginal Gyrus, anterior division Left | 0.424279663 | 0.089623943 | -0.016019866 | 0.951340214 | 0.533415183 | 0.027448263 |
| pSMG r (Supramarginal Gyrus, posterior division Ri | 0.106683037 | 0.68362067 | 0.457224535 | 0.065002018 | 0.268546954 | 0.29731395 |
| pSMG l (Supramarginal Gyrus, posterior division Le | 0.351931397 | 0.165953884 | 0.149550854 | 0.566723028 | 0.496627929 | 0.042571778 |
| AG r (Angular Gyrus Right) | 0.051502156 | 0.844372606 | 0.377326116 | 0.135412427 | 0.193746204 | 0.45621544 |
| AG l (Angular Gyrus Left) | -0.003678725 | 0.988820113 | 0.098890235 | 0.70572364 | 0.318822868 | 0.212295578 |
| sLOC r (Lateral Occipital Cortex, superior division Ri | -0.307766992 | 0.229431984 | 0.244734676 | 0.34378536 | 0.213366073 | 0.410932019 |
| sLOC l (Lateral Occipital Cortex, superior division Le | -0.04782343 | 0.855374717 | 0.235217398 | 0.363456155 | 0.307786692 | 0.229431984 |
| iLOC r (Lateral Occipital Cortex, inferior division Rig | -0.174126336 | 0.503883982 | 0.170285983 | 0.513481121 | 0.496627929 | 0.042571778 |
| iLOC l (Lateral Occipital Cortex, inferior division Lef | -0.024524836 | 0.925562269 | 0.243050527 | 0.347221056 | 0.387492409 | 0.124355518 |
| ICC r (Intracalcarine Cortex Right) | -0.34089522 | 0.180566167 | 0.577790398 | 0.015135382 | 0.289393065 | 0.259897822 |
| ICC l (Intracalcarine Cortex Left) | -0.240343393 | 0.352784473 | 0.409454808 | 0.10265421 | 0.414469728 | 0.098104229 |
| MedFC (Frontal Medial Cortex) | -0.475781818 | 0.053555409 | 0.252788164 | 0.327626709 | 0.241569635 | 0.350258203 |
| SMA r (Juxtapositional Lobule Cortex -formerly Sup | 0.209687348 | 0.419234893 | 0.656780627 | 0.004181145 | 0.186388754 | 0.473819188 |
| SMA L(Juxtapositional Lobule Cortex -formerly Supp | 0.144696532 | 0.579519874 | 0.441366434 | 0.076135405 | 0.431637114 | 0.08362004 |
| SubCalC (Subcallosal Cortex) | -0.285714339 | 0.26627772 | 0.385671917 | 0.12628751 | 0.347026429 | 0.172346387 |
| PaCiG r (Paracingulate Gyrus Right) | -0.062538332 | 0.811536581 | 0.02136287 | 0.935139085 | 0.262415745 | 0.308901139 |
| PaCiG l (Paracingulate Gyrus Left) | 0.110361762 | 0.673269147 | 0.002483103 | 0.992453568 | 0.469650609 | 0.057156103 |
| AC (Cingulate Gyrus, anterior division) | 0.3813612 | 0.130945457 | -0.278362342 | 0.279314556 | 0.480686786 | 0.050799101 |
| PC (Cingulate Gyrus, posterior division) | 0.017167385 | 0.947858964 | 0.16607779 | 0.52409436 | 0.389944892 | 0.121785592 |
| Precuneous (Precuneous Cortex) | -0.066217057 | 0.800654217 | 0.008049883 | 0.975538597 | 0.34089522 | 0.180566167 |
| Cuneal r (Cuneal Cortex Right) | -0.454935708 | 0.066529761 | 0.192715999 | 0.45866031 | 0.385039925 | 0.129963076 |
| Cuneal l (Cuneal Cortex Left) | -0.213366073 | 0.410932019 | 0.1070427 | 0.682606228 | 0.508890347 | 0.036962294 |
| FOrb r (Frontal Orbital Cortex Right) | -0.263641987 | 0.306562607 | 0.705828394 | 0.001545207 | 0.165542643 | 0.5254512 |
| FOrb l (Frontal Orbital Cortex Left) | -0.175352577 | 0.500837555 | 0.243643266 | 0.346009634 | 0.305334208 | 0.233356236 |
| aPaHC r (Parahippocampal Gyrus, anterior division | -0.294298032 | 0.251540063 | 0.146643385 | 0.574373054 | 0.468424368 | 0.057897343 |
| aPaHC l (Parahippocampal Gyrus, anterior division | -0.35438388 | 0.16281833 | 0.432536371 | 0.082906743 | 0.050275914 | 0.848037054 |
| pPaHC r (Parahippocampal Gyrus, posterior divisior | -0.339668979 | 0.182240852 | 0.342559462 | 0.17830972 | 0.147149016 | 0.573039516 |
| pPaHC l (Parahippocampal Gyrus, posterior division | -0.128755389 | 0.622370097 | 0.214468198 | 0.408461682 | 0.485591753 | 0.048150499 |
| LG r (Lingual Gyrus Right) | -0.312691659 | 0.221710351 | 0.09261199 | 0.723695044 | 0.562844986 | 0.018658872 |

**Static functional connectivity (sFC)**

| Region | clustering coefficient (CC): Kendall's tau | p-value | average path length(APL): Kendall's tau | p-value | betweenness centrality (BC): Kendall's tau | p-value |
|---|---|---|---|---|---|---|
| Frontal Pole Right | -0.412269939 | 0.100081959 | -0.007995104 | 0.975705008 | 0.241569635 | 0.350258203 |
| Frontal Pole Left | 0.275904405 | 0.283758178 | 0.175675808 | 0.500035992 | -0.316370384 | 0.21602991 |
| Insular Cortex Right | -0.206008622 | 0.427625313 | 0.060923365 | 0.816324402 | 0.091968135 | 0.725545991 |
| Insular Cortex Left | -0.345800188 | 0.173969914 | -0.116779547 | 0.655343208 | 0.369098782 | 0.144844431 |
| Superior Frontal Gyrus Right | 0.161863918 | 0.53482175 | 0.449848283 | 0.070020066 | -0.182710028 | 0.482744606 |
| Superior Frontal Gyrus Left | -0.015941143 | 0.951579066 | 0.249693252 | 0.333783302 | -0.099325586 | 0.704482768 |
| Middle Frontal Gyrus Right | -0.133660356 | 0.609054423 | -0.10747623 | 0.681384128 | 0.040465979 | 0.877452788 |
| Middle Frontal Gyrus Left | -0.143470291 | 0.582771555 | 0.217311275 | 0.402125843 | 0.071122024 | 0.786198469 |
| Inferior Frontal Gyrus, pars triangularis Right | 0.051502156 | 0.844372606 | 0.174340119 | 0.503352238 | -0.244022118 | 0.345236611 |
| Inferior Frontal Gyrus, pars triangularis Left | -0.038013496 | 0.884831959 | 0.101236497 | 0.699044498 | -0.164316401 | 0.528566334 |
| Inferior Frontal Gyrus, pars opercularis Right | 0.215818557 | 0.405445776 | 0.003694606 | 0.988771852 | -0.09564686 | 0.71499007 |
| IFG oper l (Inferior Frontal Gyrus, pars opercularis L | -0.052728397 | 0.840711173 | -0.136196319 | 0.602214868 | -0.014714902 | 0.955300113 |
| PreCG r (Precentral Gyrus Right) | -0.171673852 | 0.510002995 | -0.077827615 | 0.766543202 | 0.203556139 | 0.433267183 |
| PreCG l (Precentral Gyrus Left) | -0.202329897 | 0.436102506 | 0.286163618 | 0.265493423 | 0.186388754 | 0.473819188 |
| TP r (Temporal Pole Right) | -0.007357451 | 0.977642192 | 0.342331288 | 0.178617971 | -0.182710028 | 0.482744606 |
| TP l (Temporal Pole Left) | -0.300429241 | 0.241331849 | 0.200369118 | 0.440656064 | 0.215818557 | 0.405445776 |
| aSTG r (Superior Temporal Gyrus, anterior division | -0.283261856 | 0.270584117 | 0.142418502 | 0.585566691 | 0.023298594 | 0.929275176 |
| aSTG l (Superior Temporal Gyrus, anterior division | -0.241569635 | 0.350258203 | 0.009213766 | 0.972003091 | -0.011036176 | 0.966468204 |
| pSTG r (Superior Temporal Gyrus, posterior divisior | -0.014714902 | 0.955300113 | 0.194342531 | 0.454803251 | -0.121397938 | 0.64255182 |
| pSTG l (Superior Temporal Gyrus, posterior division | 0.101778069 | 0.697505734 | 0.219422623 | 0.397455071 | -0.171673852 | 0.510002995 |
| aMTG r (Middle Temporal Gyrus, anterior division R | -0.328632802 | 0.197777552 | 0.004907975 | 0.985084683 | 0.118945455 | 0.649332825 |
| aMTG l (Middle Temporal Gyrus, anterior division L | -0.219497282 | 0.397290445 | -0.025184294 | 0.923566108 | 0.045370947 | 0.862723602 |
| pMTG r (Middle Temporal Gyrus, posterior division | -0.111588004 | 0.669830818 | 0.3035735 | 0.236199685 | -0.186388754 | 0.473819188 |
| pMTG l (Middle Temporal Gyrus, posterior division | 0.049049672 | 0.851704448 | 0.04782343 | 0.855374717 | -0.030656045 | 0.907020959 |
| toMTG r (Middle Temporal Gyrus, temporooccipita | -0.067443299 | 0.797034351 | -0.136893826 | 0.600339129 | 0.19742493 | 0.447538998 |
| toMTG l (Middle Temporal Gyrus, temporooccipita | -0.052728397 | 0.840711173 | 0.155693045 | 0.550707693 | 0.035561012 | 0.892220091 |
| aITG r (Inferior Temporal Gyrus, anterior division Ri | -0.220858896 | 0.394294543 | 0.09864485 | 0.706423367 | 0.17044761 | 0.5130755 |
| aITG l (Inferior Temporal Gyrus, anterior division Le | -0.209687348 | 0.419234893 | 0.060122699 | 0.818700391 | 0.079705717 | 0.76106166 |
| pITG r (Inferior Temporal Gyrus, posterior division R | -0.071122024 | 0.786198469 | 0.38906028 | 0.122708241 | -0.228080975 | 0.378610298 |
| pITG l (Inferior Temporal Gyrus, posterior division L | 0.08828941 | 0.736148838 | 0.149292862 | 0.567400071 | -0.134886598 | 0.605743349 |
| toITG r (Inferior Temporal Gyrus, temporooccipital | -0.296750516 | 0.247424926 | 0.224402249 | 0.386555922 | -0.137339082 | 0.599142984 |
| toITG l (Inferior Temporal Gyrus, temporooccipital | -0.131207873 | 0.61569809 | -0.069981597 | 0.789553789 | 0.098099344 | 0.707979696 |
| PostCG r (Postcentral Gyrus Right) | -0.190067479 | 0.464975812 | 0.214022788 | 0.409459083 | 0.188841237 | 0.467914428 |
| PostCG l (Postcentral Gyrus Left) | -0.435315839 | 0.080729949 | 0.142944785 | 0.584167392 | 0.267320712 | 0.299610278 |
| SPL r (Superior Parietal Lobule Right) | -0.389944892 | 0.121785592 | -0.014162658 | 0.956976193 | 0.418148454 | 0.094859552 |
| SPL l (Superior Parietal Lobule Left) | -0.21827104 | 0.399998983 | 0.064079462 | 0.806973537 | 0.310239175 | 0.22555004 |
| aSMG r (Supramarginal Gyrus, anterior division Rig | -0.274678163 | 0.285990992 | 0.105101592 | 0.688087233 | -0.040465979 | 0.877452788 |
| aSMG l (Supramarginal Gyrus, anterior division Left | -0.060085848 | 0.818809784 | 0.239705386 | 0.354102949 | -0.1496015 | 0.566590159 |
| pSMG r (Supramarginal Gyrus, posterior division Ri | -0.123850422 | 0.6357974 | -0.253998309 | 0.325237494 | 0.172900094 | 0.506939142 |
| pSMG l (Supramarginal Gyrus, posterior division Le | -0.060085848 | 0.818809784 | 0.110429448 | 0.673079198 | 0.042918463 | 0.870083148 |
| AG r (Angular Gyrus Right) | -0.328632802 | 0.197777552 | -0.042971156 | 0.869924915 | 0.289393065 | 0.259897822 |
| AG l (Angular Gyrus Left) | -0.056407123 | 0.829745661 | -0.016564417 | 0.949688091 | 0.050275914 | 0.848037054 |
| sLOC r (Lateral Occipital Cortex, superior division Ri | 0.07480075 | 0.775399567 | 0.154982019 | 0.552551377 | -0.101778069 | 0.697505734 |
| sLOC l (Lateral Occipital Cortex, superior division Le | -0.187614995 | 0.470862232 | 0.052825593 | 0.840421089 | 0.025751078 | 0.921850815 |
| iLOC r (Lateral Occipital Cortex, inferior division Rig | -0.414469728 | 0.098104229 | 0.064456734 | 0.805857386 | 0.147149016 | 0.573039516 |
| iLOC l (Lateral Occipital Cortex, inferior division Lef | -0.107909278 | 0.680164132 | 0.289589878 | 0.259559191 | 0.164316401 | 0.528566334 |
| ICC r (Intracalcarine Cortex Right) | 0.019619869 | 0.940421887 | 0.377847942 | 0.134828896 | -0.082158201 | 0.753919876 |
| ICC l (Intracalcarine Cortex Left) | -0.177805061 | 0.49477106 | 0.023327199 | 0.929188548 | 0.32004911 | 0.210444172 |
| MedFC (Frontal Medial Cortex) | -0.265030675 | 0.303927005 | 0.029648615 | 0.910064644 | -0.02697732 | 0.918140885 |
| SMA r (Juxtapositional Lobule Cortex -formerly Sup | -0.206008622 | 0.427625313 | 0.007361963 | 0.977628483 | 0.361741331 | 0.153652534 |
| SMA L(Juxtapositional Lobule Cortex -formerly Supp | -0.160637676 | 0.537961945 | 0.185276074 | 0.476510191 | -0.019619869 | 0.940421887 |
| SubCalC (Subcallosal Cortex) | 0.338442737 | 0.18392582 | 0.023941073 | 0.927329655 | -0.404659794 | 0.107143314 |
| PaCiG r (Paracingulate Gyrus Right) | -0.150827741 | 0.563377227 | -0.019631902 | 0.940385408 | -0.014714902 | 0.955300113 |
| PaCiG l (Paracingulate Gyrus Left) | -0.068669541 | 0.793418393 | -0.026710136 | 0.918949103 | -0.111588004 | 0.669830818 |
| AC (Cingulate Gyrus, anterior division) | -0.029429803 | 0.910725885 | -0.08067485 | 0.758987145 | 0.19742493 | 0.447538998 |
| PC (Cingulate Gyrus, posterior division) | -0.160637676 | 0.537961945 | -0.087277346 | 0.739073819 | 0.277130647 | 0.281535969 |
| Precuneous (Precuneous Cortex) | -0.295524274 | 0.249477184 | -0.186617592 | 0.473266675 | 0.41569597 | 0.097013982 |
| Cuneal r (Cuneal Cortex Right) | 0.112814246 | 0.666398678 | 0.251533742 | 0.330114077 | -0.242795876 | 0.347742244 |
| Cuneal l (Cuneal Cortex Left) | -0.077253233 | 0.768221732 | 0.086769642 | 0.740542421 | -0.012262418 | 0.962744755 |
| FOrb r (Frontal Orbital Cortex Right) | -0.011036176 | 0.966468204 | 0.234173723 | 0.365650877 | -0.042918463 | 0.870083148 |
| FOrb l (Frontal Orbital Cortex Left) | -0.217044799 | 0.402717435 | 0.224616448 | 0.386090796 | 0.038013496 | 0.884831959 |
| aPaHC r (Parahippocampal Gyrus, anterior division | -0.062538332 | 0.811536581 | 0.276138597 | 0.283332954 | -0.040465979 | 0.877452788 |
| aPaHC l (Parahippocampal Gyrus, anterior division | -0.062538332 | 0.811536581 | 0.117321956 | 0.65383613 | 0.117719213 | 0.652733157 |
| pPaHC r (Parahippocampal Gyrus, posterior divisior | -0.087063168 | 0.739693253 | -0.023370304 | 0.929058008 | 0.080931959 | 0.757488449 |
| pPaHC l (Parahippocampal Gyrus, posterior division | -0.10913552 | 0.676713604 | 0.089563525 | 0.732471368 | -0.002452484 | 0.99254662 |
| LG r (Lingual Gyrus Right) | -0.248927085 | 0.335317659 | 0.059582355 | 0.820304719 | -0.003678725 | 0.988820113 |

| | | | | | | | | | | | | | | | | | | |
|---|---|---|---|---|---|---|---|---|---|---|---|---|---|---|---|---|---|---|
| LG l (Lingual Gyrus Left) | -0.347026429 | 0.172346387 | 0.153509126 | 0.556379181 | -0.241569635 | 0.350258203 | -0.175352577 | 0.500837555 | 0.172654994 | 0.507550847 | 0.380134958 | 0.132291971 | -0.067443299 | 0.797034351 | 0.233703044 | 0.366643077 | -0.082158201 | 0.753919876 |
| aTFusC r (Temporal Fusiform Cortex, anterior divisi | -0.392397376 | 0.11925307 | 0.033210433 | 0.89930912 | 0.007357451 | 0.977642192 | -0.30656045 | 0.231388818 | 0.366707893 | 0.14766779 | 0.116492971 | 0.656139965 | 0.052728397 | 0.840711173 | -0.105651185 | 0.686533872 | 0.111588004 | 0.669830818 |
| aTFusC l (Temporal Fusiform Cortex, anterior divisi | -0.145922774 | 0.576275844 | -0.184616259 | 0.47810948 | 0.024524836 | 0.925562269 | -0.356836364 | 0.159723031 | 0.030153989 | 0.90853764 | 0.418148454 | 0.094859552 | 0.106748466 | 0.683436086 | 0.088343558 | 0.735992438 | -0.126302905 | 0.629070012 |
| pTFusC r (Temporal Fusiform Cortex, posterior divis | -0.036787254 | 0.888524941 | 0.177846996 | 0.494667637 | -0.234212184 | 0.365569868 | -0.143470291 | 0.582771555 | 0.420734431 | 0.092625245 | 0.304107966 | 0.235334242 | 0.098099344 | 0.707979696 | 0.384143247 | 0.127925891 | -0.464745642 | 0.060163967 |
| pTFusC l (Temporal Fusiform Cortex, posterior divis | -0.214592315 | 0.408183979 | 0.307825507 | 0.229370215 | -0.40220731 | 0.109492353 | -0.239117151 | 0.355321036 | 0.212217953 | 0.41351388 | 0.114040487 | 0.662972783 | 0.133660356 | 0.609054423 | -0.121456471 | 0.642390302 | 0.064990815 | 0.804277922 |
| TOFusC r (Temporal Occipital Fusiform Cortex Right | -0.42918463 | 0.085587962 | 0.047135154 | 0.857436034 | -0.299202999 | 0.243352261 | -0.452483224 | 0.068195936 | 0.598285923 | 0.01117894 | 0.190067479 | 0.464975812 | -0.213366073 | 0.410932019 | 0.233703044 | 0.366643077 | 0.150827741 | 0.563377227 |
| TOFusC l (Temporal Occipital Fusiform Cortex Left) | -0.351931397 | 0.165953884 | 0.069115276 | 0.792104985 | -0.236664667 | 0.360424972 | -0.225628491 | 0.383897317 | 0.291624164 | 0.256075089 | 0.25873702 | 0.315979873 | 0.01348866 | 0.959022034 | 0.37592166 | 0.136991626 | -0.371551265 | 0.141987036 |
| OFusG r (Occipital Fusiform Gyrus Right) | -0.546903843 | 0.023092554 | 0.472025556 | 0.05574061 | -0.105456795 | 0.687083159 | -0.311465417 | 0.223624917 | 0.411163246 | 0.101087622 | 0.360515089 | 0.155155198 | -0.142244049 | 0.586030839 | 0.086475128 | 0.74139473 | 0.196198688 | 0.450421764 |
| OFusG l (Occipital Fusiform Gyrus Left) | -0.441447048 | 0.076075508 | 0.321924601 | 0.207632813 | -0.046597188 | 0.859047792 | -0.41569597 | 0.097013982 | 0.382298835 | 0.129922297 | 0.214592315 | 0.408183979 | -0.322501593 | 0.206772833 | 0.087601597 | 0.738136325 | 0.248927085 | 0.335317659 |
| FO r (Frontal Operculum Cortex Right) | 0.187614995 | 0.470862232 | 0.141889911 | 0.586973526 | 0.208461106 | 0.42202201 | 0.028203561 | 0.914432551 | 0.094711234 | 0.717670261 | 0.365420056 | 0.149204072 | 0.036787254 | 0.888524941 | -0.02274128 | 0.930963113 | 0.056407123 | 0.829745661 |
| FO l (Frontal Operculum Cortex Left) | 0 | 1 | 0.074870259 | 0.77519589 | -0.190067479 | 0.464975812 | 0.305334208 | 0.233356236 | 0.098333419 | 0.707311745 | 0.467198126 | 0.058645707 | -0.274846626 | 0.285683618 | 0.275523589 | 0.284450456 | 0.116492971 | 0.656139965 |
| CO r (Central Opercular Cortex Right) | 0.169221368 | 0.516156614 | 0.047643873 | 0.855912387 | 0.036787254 | 0.888524941 | 0.229307217 | 0.375981928 | 0.254500142 | 0.3242497 | 0.19742493 | 0.447538998 | -0.057633365 | 0.826096984 | 0.077348081 | 0.767944489 | -0.093194377 | 0.722022052 |
| CO l (Central Opercular Cortex Left) | 0.262415745 | 0.308901139 | -0.088526429 | 0.735464319 | 0.134886598 | 0.605743349 | 0.40220731 | 0.109492353 | 0.02925004 | 0.911269163 | 0.376456233 | 0.13638905 | 0.154506467 | 0.553785992 | -0.03439806 | 0.895726464 | -0.134886598 | 0.605743349 |
| PO r (Parietal Operculum Cortex Right) | -0.284488098 | 0.268425606 | 0.175692144 | 0.499995499 | 0.286940581 | 0.264140461 | 0.274678163 | 0.285990992 | -0.077402719 | 0.767784791 | 0.475781818 | 0.053555409 | -0.217044799 | 0.402717435 | -0.00246155 | 0.992519067 | 0.350705155 | 0.167536807 |
| PO l (Parietal Operculum Cortex Left) | 0.239117151 | 0.355321036 | 0.023460143 | 0.928785946 | 0.120171696 | 0.645939027 | 0.421827179 | 0.091692562 | 0.15170933 | 0.561072212 | 0.427958388 | 0.086584387 | 0.041692221 | 0.873766741 | 0.084662577 | 0.746646392 | -0.029429803 | 0.910725885 |
| PP r (Planum Polare Right) | -0.167995127 | 0.519246297 | 0.083352249 | 0.750449508 | 0.087063168 | 0.739693253 | -0.147149016 | 0.573039516 | 0.306086911 | 0.232147322 | 0.408338519 | 0.103687105 | 0.042918463 | 0.870083148 | -0.227747127 | 0.379327629 | 0.04782343 | 0.855374717 |
| PP l (Planum Polare Left) | 0.045370947 | 0.862723602 | 0.019891831 | 0.939597447 | 0.042918463 | 0.870083148 | -0.094420619 | 0.718503392 | 0.217334356 | 0.402074623 | 0.160637676 | 0.537961945 | 0.030656045 | 0.907020959 | 0.038130497 | 0.884479712 | 0.241569635 | 0.350258203 |
| HG r (Heschl's Gyrus Right) | -0.112814246 | 0.666398678 | -0.011503864 | 0.965048002 | -0.049049672 | 0.851704448 | 0.191293721 | 0.462046419 | 0.426198843 | 0.088028774 | 0.360515089 | 0.155155198 | -0.153280225 | 0.556975096 | -0.176795613 | 0.497263773 | 0.378908716 | 0.133648057 |
| HG l (Heschl's Gyrus Left) | -0.365420056 | 0.149204072 | 0.065683517 | 0.80223043 | -0.213366073 | 0.410932019 | -0.35438388 | 0.16281833 | 0.222364614 | 0.390995881 | 0.423053421 | 0.090654005 | -0.018393627 | 0.94413988 | -0.000613874 | 0.998134345 | 0.198651172 | 0.444665657 |
| PT r (Planum Temporale Right) | 0.085836926 | 0.743242629 | -0.037622334 | 0.886009752 | -0.036787254 | 0.888524941 | 0.29307179 | 0.253613565 | 0.363648116 | 0.151335647 | 0.29307179 | 0.253613565 | 0.21827104 | 0.399998983 | -0.144785276 | 0.579284845 | -0.029429803 | 0.910725885 |
| PT l (Planum Temporale Left) | 0.185162512 | 0.47678526 | -0.127530716 | 0.625712306 | 0.106683037 | 0.68362067 | 0.17044761 | 0.5130755 | 0.186423305 | 0.473735746 | 0.447578257 | 0.071620089 | -0.226854733 | 0.381248768 | 0.282903685 | 0.271216597 | -0.24524836 | 0.342741321 |
| SCC r (Supracalcarine Cortex Right) | -0.421827179 | 0.091692562 | -0.097898243 | 0.708553714 | 0.267320712 | 0.299610278 | -0.554261294 | 0.020954638 | 0.716204331 | 0.001220768 | 0.103004311 | 0.694025752 | 0.07480075 | 0.775399567 | 0.416617357 | 0.096200507 | -0.290619307 | 0.257792443 |
| SCC l (Supracalcarine Cortex Left) | -0.085836926 | 0.743242629 | -0.141626582 | 0.587674893 | 0.377682474 | 0.135013741 | 0.052728397 | 0.840711173 | 0.303145149 | 0.236894754 | 0.62660956 | 0.007110822 | -0.042918463 | 0.870083148 | -0.006138737 | 0.981344954 | 0.069895783 | 0.78980641 |
| OP r (Occipital Pole Right) | -0.277130647 | 0.281535969 | -0.213119613 | 0.411485528 | 0.150827741 | 0.563377227 | -0.154506467 | 0.553785992 | 0.502478709 | 0.039820254 | 0.237890909 | 0.357867875 | -0.230533458 | 0.373363679 | 0.168304795 | 0.518465238 | 0.020846111 | 0.936705057 |
| OP l (Occipital Pole Left) | -0.332311528 | 0.192505433 | 0.36990432 | 0.143901583 | -0.228080975 | 0.378610298 | -0.219497282 | 0.397290445 | 0.197285287 | 0.447866808 | 0.080931959 | 0.757488449 | -0.214592315 | 0.408183979 | 0.401977762 | 0.109714068 | 0.187614995 | 0.470862232 |
| Thalamus r | -0.172900094 | 0.506939142 | 0.141624862 | 0.587679477 | 0.138565323 | 0.595853798 | -0.165542643 | 0.5254512 | 0.22160916 | 0.392649014 | 0.443899532 | 0.074269716 | -0.024524836 | 0.925562269 | 0.255843333 | 0.321614425 | -0.118945455 | 0.649332825 |
| Thalamus l | -0.111588004 | 0.669830818 | 0.101743316 | 0.697604446 | 0.185162512 | 0.47678526 | -0.362967573 | 0.152159806 | 0.465440966 | 0.059730584 | 0.166768885 | 0.522344507 | -0.121397938 | 0.64255182 | 0.09969278 | 0.703436709 | -0.031882287 | 0.903317846 |
| Caudate r | -0.226854733 | 0.381248768 | 0.04753177 | 0.856248103 | -0.079705717 | 0.76106166 | -0.430410872 | 0.084599858 | 0.494757795 | 0.0434809 | 0.117719213 | 0.652733157 | -0.132515337 | 0.612152673 | 0.422155845 | 0.091413366 | -0.045370947 | 0.862723602 |
| Caudate l | -0.053954639 | 0.837052825 | 0.185690709 | 0.475506525 | -0.053954639 | 0.837052825 | 0.176578819 | 0.497799901 | 0.040691 | 0.876776201 | 0.49049672 | 0.045607063 | -0.506437863 | 0.038036392 | 0.219152896 | 0.398050128 | -0.042918463 | 0.870083148 |
| Putamen r | -0.297976757 | 0.245383286 | 0.147775026 | 0.57139032 | -0.038013496 | 0.884831959 | -0.198651172 | 0.444665657 | 0.31038782 | 0.225316115 | 0.192519963 | 0.459126283 | 0.046597188 | 0.859047792 | 0.246474602 | 0.340256389 | -0.150827741 | 0.563377227 |
| Putamen l | -0.280809372 | 0.274933005 | 0.09212617 | 0.725091539 | 0.101778069 | 0.697505734 | -0.182710028 | 0.482744606 | 0.522365073 | 0.031467352 | 0.242795876 | 0.347742244 | 0.08828941 | 0.736148838 | 0.019124292 | 0.94192435 | 0.164316401 | 0.528566334 |
| Pallidum r | -0.488044236 | 0.046865794 | 0.539079475 | 0.025548086 | -0.324954077 | 0.203143384 | -0.240343393 | 0.352784473 | 0.442737012 | 0.075121742 | 0.160637676 | 0.537961945 | 0.015941143 | 0.951579066 | 0.117249869 | 0.654036351 | -0.099325586 | 0.704482768 |
| Pallidum l | -0.246474602 | 0.340256389 | 0.022783407 | 0.930835515 | 0.049079755 | 0.851614443 | -0.312691659 | 0.221710351 | 0.190770134 | 0.463296096 | 0.226854733 | 0.381248768 | 0.255058294 | 0.323153107 | 0.049877188 | 0.849229228 | -0.101778069 | 0.697505734 |
| Hippocampus r | -0.137339082 | 0.599142984 | 0.362577856 | 0.15263314 | -0.177805061 | 0.49477106 | -0.706315277 | 0.001528545 | 0.370764506 | 0.142899433 | 0.203556139 | 0.433267183 | 0.399631751 | 0.111998264 | 0.119631902 | 0.647432177 | -0.035561012 | 0.892220091 |
| Hippocampus l | 0.230533458 | 0.373363679 | -0.206922627 | 0.425532525 | 0.177805061 | 0.49477106 | -0.079705717 | 0.76106166 | 0.186693049 | 0.47308456 | 0.573881162 | 0.016001667 | 0.052728397 | 0.840711173 | 0.500614628 | 0.040681807 | -0.229307217 | 0.375981928 |
| Amygdala r | -0.196198688 | 0.450421764 | 0.114199707 | 0.662528415 | 0.015941143 | 0.951579066 | -0.51747404 | 0.033384043 | 0.329428096 | 0.196629859 | 0.110361762 | 0.673269147 | -0.264868229 | 0.304234614 | -0.082208589 | 0.753773337 | 0.256284536 | 0.320751547 |
| Amygdala l | -0.221949766 | 0.39190321 | 0.5043402 | 0.038973789 | -0.133660356 | 0.609054423 | -0.624157076 | 0.007407238 | 0.688047826 | 0.002264386 | 0.002452484 | 0.99254662 | -0.002452484 | 0.99254662 | 0.049200641 | 0.85125278 | 0.093194377 | 0.722022052 |
| Accumbens r | -0.019619869 | 0.940421887 | 0.146986915 | 0.573466895 | 0.057633365 | 0.826096984 | -0.128755389 | 0.622370097 | 0.427785928 | 0.086725195 | 0.219497282 | 0.397290445 | 0.418148454 | 0.094859552 | -0.009236516 | 0.971933988 | -0.229307217 | 0.375981928 |
| Accumbens l | -0.120171696 | 0.645939027 | 0.185806009 | 0.475227614 | 0.203556139 | 0.433267183 | -0.137339082 | 0.599142984 | 0.231528674 | 0.371246169 | 0.252605811 | 0.327987614 | -0.301655483 | 0.239322044 | -0.066257669 | 0.80053427 | 0.163090159 | 0.531689865 |
